# Mapping the brain signature of dyslexia risk onto histamine, acetylcholine and glutamate receptor systems

**DOI:** 10.64898/2026.10.18.26365830

**Authors:** Murat Can Çelik, Yusuf Çiçek, Mehmet Akif Şakiroğlu, Doğukan Aslan, Ali Tarık Altunç, Halil Aziz Velioğlu

## Abstract

The brain differences of dyslexia are well described and its genetic risk is now measurable; the brain systems that connect the two are unknown. That link decides which measures complement one another in early identification and which systems intervention research should examine. Here we compared large-scale polygenic score maps of brain structure in dyslexia with normative maps of brain neurotransmitter systems. The structural signature of genetic liability aligned with the cortical distribution of neurotransmitter systems. The strongest and most consistent alignment appeared for the histamine H3 receptor, the vesicular acetylcholine transporter (VAChT) and the N-methyl-D-aspartate (NMDA) glutamate receptor. The alignment was most pronounced in cingulate, medial and superior frontal cortex and the paracentral lobule. The H3 alignment held after adjustment for cortical position and for metabolic and cognitive reference maps. Once the component common to all nineteen maps and position were removed together, no individual system remained significant, while the shared component itself aligned with liability. The signature therefore follows the topography these systems share. In 169 children, per-person alignment with the H3 map related to reading skill with the opposite sign to liability. To our knowledge, this is the first study to relate the structural signature of genetic liability for dyslexia to normative maps of neurotransmitter receptor and transporter density. It places that signature within the shared geography of the systems that set attention, arousal and synaptic plasticity during learning, and gives the genetics of dyslexia a testable biological axis.

## Introduction

Dyslexia is the most common specific learning disorder [1, 2]. It emerges in the school years, and its consequences reach into education and working life [1]. It is also strongly heritable [3]. These two properties make dyslexia a natural model for studying how genetic liability shapes the brain [4].

Functional studies of diagnosed readers converge on a left-hemisphere reading network, with occipitotemporal, temporoparietal and inferior frontal nodes that are underactivated in dyslexia across languages [5]. Structural differences cluster in the same perisylvian and occipitotemporal territory, although meta-analyses and large samples show many inconsistencies [6, 7]. These maps describe the outcome, the brain of a person who already reads with difficulty, and part of what they show follows from living with the disorder [7].

On the genetic side, a study of 51 800 diagnosed adults identified 42 genomic loci for dyslexia [3]. On the brain side, the polygenic score, a single number summarising the genetic liability a person carries, was related to brain structure in more than 35 000 adults [4]. The clearest signature of liability ran along the medial wall, from parietal to frontal cortex [4]. Where liability marks the brain is therefore known [4, 8].

Location alone leaves the biology open, because a mapped signature says where liability acts and leaves the tissue property behind it unresolved [4]. The same cortical territory can be marked through metabolic, hierarchical or neurochemical routes, and each route means something different [9, 10]. It names a candidate mechanism, because neuromodulators set the dynamics on which learning runs [11]. Receptors and transporters shape signalling between neurons, and direct density measures capture them more faithfully than gene expression [12].

Neurotransmitter systems are the strongest candidate for such an answer [13]. They are laid out across the cortex in an orderly way, and they set attention, arousal and synaptic plasticity during learning [11]. Nineteen receptor and transporter targets across nine of these systems are mapped in an open normative positron emission tomography (PET) atlas [13]. The atlas has been related to brain maps of psychiatric and neurological disorders [13]. Polygenic risk maps have been carried to brain structure in psychiatry [14] and in childhood epilepsy [15]. Reading and mathematical skill in typically developing children have been placed in this literature [16]. Polygenic risk has been related to receptor maps in Alzheimer’s disease [17], and the genetics of word reading has been located in cortical cell types, without a brain map [18]. Genetic liability for dyslexia has yet to be placed on a receptor atlas [17, 18]. The authors of the source study explicitly called for receptor-level examination of the affected regions, and this study takes that step [4].

Brain maps are spatially smooth, so comparisons of this kind can produce misleading alignment [19]. We therefore grounded the comparison in stringent statistical tests that account for spatial structure and in a control family of cognitive and metabolic reference maps [13, 19].

In this study we compared the brain signature of genetic liability for dyslexia [4] with nineteen neurotransmitter receptor and transporter maps across the cortex [13]. We asked in which systems alignment emerges, where in the cortex it concentrates, and whether it separates from position, hierarchy and metabolism [9, 10].

## Materials and methods

Maps relating polygenic liability for dyslexia to brain structure came from the open archive of Soheili-Nezhad et al. [4]; Table 1 summarises every data source. Polygenic scores in that study derive from the genome-wide association study (GWAS) of Doust et al. [3], which identified 42 loci in 51 800 adults who reported a dyslexia diagnosis and 1 087 070 controls. Scores were related to tensor-based morphometry in up to 35 231 unrelated white British adults aged 45 to 82 years. We used the released pooled-sex, unthresholded voxel-wise t maps in three constructions. These were unadjusted log-Jacobians, raw Jacobians adjusted for head size, and log-Jacobians adjusted for head size, education and verbal-numerical reasoning. Maps were brought to Montreal Neurological Institute (MNI152) space with the released Advanced Normalization Tools (ANTs) transforms [20]. They were averaged within the 218 Brainnetome regions used by Zhang et al. [16], 210 cortical parcels plus the amygdala and hippocampal subregions [21], and standardised. The three constructions correlated at r = 0.90 to 0.98. The third adjusts for education and verbal-numerical reasoning, which may lie downstream of liability, so the three are alternative specifications, and the third gives the smallest correlations for VAChT and NMDA.

**Table 1.** Data sources.

| Component | Source | Sample | Measure | Regions |
| --- | --- | --- | --- | --- |
| Liability maps | Soheili-Nezhad et al. [4]; polygenic scores from Doust et al. [3] | 35 231 UK Biobank adults, 45 to 82 years | Tensor-based morphometry t maps; three covariate constructions | 218 Brainnetome regions |
| Neurotransmitter maps | Hansen et al. [13] as distributed by Zhang et al. [16] | PET group averages, more than 1 200 healthy adults (8 to 204 per system; Supplementary Table 1) | Nineteen receptor and transporter densities | 218 Brainnetome regions |
| Reference maps | neuromaps [27] | Meta-analytic and group-average maps | Neurosynth cognitive component; cerebral blood flow; glucose metabolism | 218 Brainnetome regions |
| Reading-skill map, cohort 1 | OpenNeuro ds001894 [30] | 169 children, 7 to 14 years | TOWRE score against log-Jacobian morphometry | 218 Brainnetome regions |
| Reading-skill map, cohort 2 | Carrion-Castillo et al. [31], published tables | 9 013 ABCD children, 9 to 10 years | Reading performance against surface area and thickness | 74 left-hemisphere Destrieux regions |

Nineteen receptor and transporter maps came from the PET atlas of Hansen et al. [13] as parcellated by Zhang et al. [16], with the corrected VAChT map. Source samples range from 8 participants for H3 to 204 for MOR, and VAChT combines three studies of 4, 5 and 18 participants (Supplementary Table 1). The NMDA map derives from [18F]GE-179, which binds the open channel and indexes activated receptors more than total density [22, 23]. The systems were the serotonin receptors 5-HT1A, 5-HT1B, 5-HT2A, 5-HT4 and 5-HT6, the serotonin transporter (5-HTT), nicotinic α4β2, cannabinoid CB1, dopamine D1 and D2, the dopamine transporter (DAT), GABAA, histamine H3, muscarinic M1, mGluR5, the mu-opioid receptor (MOR), the noradrenaline transporter (NET), the N-methyl-D-aspartate (NMDA) receptor and VAChT. Maps were averaged within the same 218 regions and standardised. The analysis chain reproduced the 38 normative reading-receptor coefficients published by Zhang et al. [16] from the same inputs.

Each liability map was correlated (Pearson) with each receptor map across the 218 regions (Figure 1c). Because brain maps are spatially autocorrelated, probabilities came from 5 000 surrogate maps that preserve the spatial structure of the target, generated by three procedures. Moran spectral randomisation [24] preserves global autocorrelation by flipping the signs of the target’s projections onto the eigenvectors of a double-centred inverse-distance weight matrix. Variogram-matched surrogates (BrainSMASH) [19] preserve distance-dependent autocorrelation by smoothing permuted maps until their variogram matches the target’s. A distribution-matched variant additionally resamples surrogate values from the target’s own distribution. Distances were Euclidean distances between region centroids in MNI space. Two-sided P equalled (1 + number of surrogate correlations at least as extreme)/5 001, and the Benjamini-Hochberg procedure controlled the false discovery rate across the 57 tests within each procedure [25]. A map counted as aligned when it reached q < 0.05 with the same sign in all three constructions under all three procedures. Later analyses reuse this inference (Supplementary Table 2). The constructions correlate at r = 0.90 to 0.98 and the two variogram-matched procedures agree closely, so the nine cells of a test are a sensitivity check on one dataset. Back-solving the effective n from each observed correlation and its surrogate probability gave a median of 39 (interquartile range 27 to 59; Supplementary Table 3). Comparisons between correlations are therefore reported at the nominal n of 218 and, as an approximate sensitivity calculation, at an effective n of 39.

**Figure 1.**
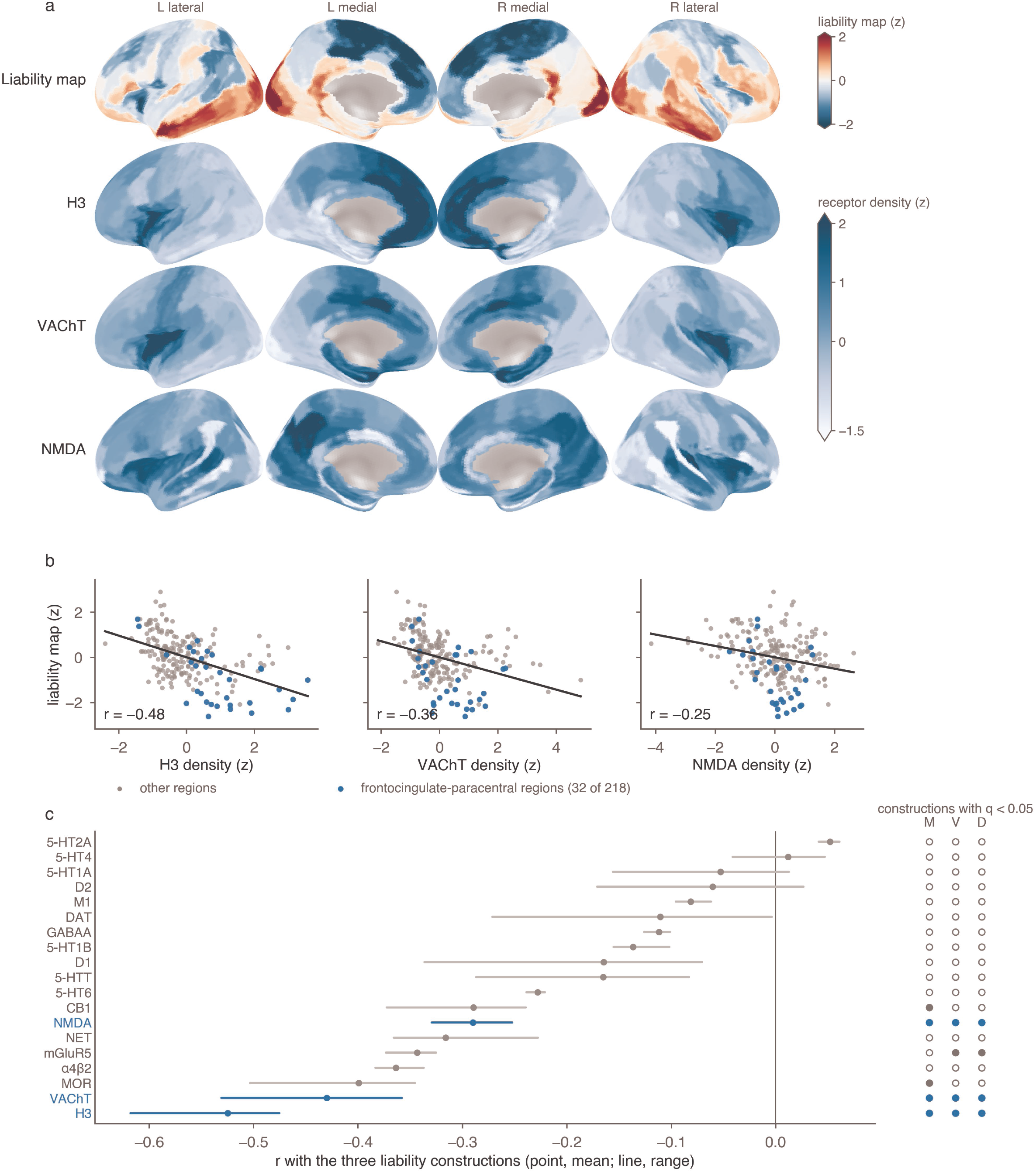
Genetic liability for dyslexia follows neurotransmitter topography. (a) The education-adjusted liability map and the normative H3, VAChT and NMDA densities on the inflated fsaverage5 surface, in lateral and medial views of both hemispheres; column headers give hemisphere and view. All maps are standardised across the 218 regions, and negative liability values mark lower regional volume with higher polygenic liability. The upper colour bar (diverging) applies to the liability map and the lower (sequential) to the three receptor maps; the two scales differ, and the tapered ends mark the parcels that fall outside them (liability 14, H3 16, VAChT 14 and NMDA 16 of 218). (b) Region-wise relation between each of the three receptor maps and the liability map across the 218 regions, with the 32 frontocingulate-paracentral regions in blue and the least-squares line in black. Panel b shows the education-adjusted construction, the most adjusted of the three, deliberately; it gives the smallest correlation of the three for VAChT and NMDA and comes within 0.01 of the smallest for H3. (c) Correlation of each of the nineteen maps with the three liability constructions (point, mean; line, range). The three systems that reached q < 0.05 in all three constructions under all three procedures are in blue. The marks to the right lie on their own scale, not on the correlation axis, and give the number of constructions reaching q < 0.05 under Moran (M), variogram-matched (V) and distribution-matched (D) surrogates: solid, all three constructions; open, none. No system falls between the two. Mark colour follows the system, blue for the three retained maps and grey for the rest.

For the position control, target and receptor maps were residualised on nine geometric terms computed at region centroids. These were the standardised x, y and z coordinates, their squares and their pairwise products. The same adjustment was applied to the normative child reading-structure maps of Zhang et al. [16], correlated with the same targets, as a positional comparison. A further control removed each region’s fractional membership in the 14 resting-state networks of Shirer et al. [26] (Supplementary Methods).

Three non-molecular cortical maps came from neuromaps [27] as MNI152 volumes and were parcellated identically. They were the first principal component of Neurosynth activation maps [28], mean cerebral blood flow [29] and the cerebral metabolic rate of glucose [9]. They served first as substitute targets and then as simultaneous covariates. For each receptor, a leave-one-out common axis was computed as the first principal component of the other eighteen maps, so that the map under test never entered its own covariate. The screen was repeated with this component, with the nine geometric terms, and with both.

The frontocingulate-paracentral territory was defined before analysis by Brainnetome label prefixes as the superior frontal, cingulate and paracentral parcels (32 regions, 14.7%). Its contribution was the share of the standardised cross-product sum carried by these regions. The prespecified H3, NMDA and VAChT tests were repeated with the territory removed (186 regions) and within it alone, with distance matrices restricted accordingly. Sensitivity analyses added the insula to the removed set (44 regions), removed the entire frontal lobe with cingulate, insula and paracentral cortex (82 regions), and removed both poles of the gradient (68 regions).

Reading-skill maps were built from OpenNeuro ds001894 (188 children aged 7.36 to 14.54 years) [30]. Each image was registered to the study template with affine plus symmetric normalisation (SyN) transformations [20]. The primary measure was the geometric log-Jacobian of the composed field, matching the adult maps, and the nonlinear-only log-Jacobian served as a sensitivity definition. Warped-template correlation was bimodal, with 169 scans between 0.633 and 0.721 and 19 scans between -0.150 and 0.525 and none in between. The 19 were excluded, leaving 169 children (mean age 10.70 years, SD 1.64; Test of Word Reading Efficiency (TOWRE) total 96.1, SD 17.7), and a stricter threshold of 0.66 (152 children) served as a sensitivity analysis. Parcel-wise linear models with heteroscedasticity-consistent (HC3) standard errors related the standardised TOWRE score to the standardised regional log-Jacobian, adjusting for age, sex and mean brain log-Jacobian. The resulting 218-region t map was screened against the nineteen receptor maps, with false discovery control across the 19 maps and across the three prespecified maps. Its similarity to each adult liability map was tested in the same way. Sensitivity models added quadratic age, full-scale IQ, magnetic resonance imaging (MRI) quality metrics, removal of registration and image-quality outliers, omission of the global covariate and an alternative reading measure. The adult and child H3 correlations were compared with the Williams test. The child map’s reliability, estimated from the regional standard errors, served to disattenuate its correlation and to compute power. A prespecified per-person analysis defined each child’s deviation vector as the regional log-Jacobians residualised across children on age, sex and mean brain log-Jacobian. Each vector was correlated with the H3, VAChT and NMDA maps and the shared axis. Reading skill was then regressed on each score with the same covariates.

A second skill map came from the published tables of Carrion-Castillo et al. [31], which related reading performance to surface area and thickness in 9 013 children of the Adolescent Brain Cognitive Development (ABCD) Study across the 74 left-hemisphere regions of the Destrieux atlas [32]. Models fitted with adjustment for total surface area or mean thickness served as primary maps and the unadjusted models as sensitivity maps.

The nineteen receptor maps were rebuilt at the same 74 regions. Each MNI152 receptor volume was projected to the fsaverage surface with the registration-fusion transform in neuromaps and averaged within Destrieux labels. Source studies were combined with the weights of Hansen et al. [13], and distances were Euclidean distances between region centroids on the fsaverage pial surface. False discovery control ran across 38 tests (19 maps x 2 measures) and across the six prespecified tests. Similarity between each skill map and the three adult liability maps, re-parcellated into the same 74 regions, was tested in the same way.

Analyses used Python 3.9 with numpy, pandas, scipy, nibabel, neuromaps and brainsmash 0.11.0, with fixed seeds. Secondary analyses, comprising case-control and task-based functional magnetic resonance imaging (fMRI) representations of dyslexia, source-map uncertainty and a summary-statistics genetic comparison of word-reading and cortical GWAS [33–35], used the same framework. The territory analysis, the comparison with dyslexia maps, a stratified phenotype family, the two published-map families and the per-person analysis were prespecified in dated documents. The primary screen and the three control families have no separate document. In total the study comprises 26 test families, 745 tests and 1 842 probabilities in 91 false discovery families (Supplementary Table 2).

## Results

Across the nineteen receptor maps, the liability maps aligned with a small and consistent set of systems (Figure 1a-c; Supplementary Table 4). Under Moran randomisation, five maps were inversely associated with all three constructions, and requiring agreement under both variogram-matched procedures reduced this set to H3, VAChT and NMDA (Figure 1c). H3 correlated with the three constructions at r = -0.618 to -0.475, NMDA at -0.329 to -0.252 and VAChT at -0.531 to -0.358 (all P <= 0.0054; worst q = 0.0249). CB1 and MOR passed under Moran randomisation only, and mGluR5 under the variogram-matched procedures only. The negative sign indicates lower regional volume with higher polygenic liability where normative receptor density is high. The architecture the nineteen maps share aligned with liability directly. Their first principal component, carrying 38% of receptor variance, correlated with the three constructions at r = -0.43 to - 0.30 (Supplementary Table 5). It reached q < 0.05 in all three under both variogram-matched procedures and in two under Moran randomisation. The three retained maps are not the strongest loadings on this component (H3 ranks 6, NMDA 9 and VAChT 14 of nineteen; Supplementary Table 6).

Ranking regions by the product of standardised H3 density and liability effect placed the alignment in cingulate cortex, superior frontal cortex, insula and the paracentral lobule (Figure 1a,b). By gyral grouping, H3 density was highest in cingulate (1.13) and prefrontal cortex (0.75) and lowest in occipital (-0.98) and inferior temporal cortex (-0.74). The liability effect followed the same axis, most negative in the paracentral lobule (-2.04) and cingulate cortex (-0.53) and positive in occipital (1.11) and inferior temporal cortex (0.82; Supplementary Table 7).

The frontocingulate-paracentral territory, 32 of 218 regions, carried 40 to 50% of the H3 cross-product sum and 20 to 30% of the NMDA and VAChT sums. With the territory removed, H3, NMDA and VAChT remained significant in every construction under every procedure (H3 r = -0.525, -0.371 and -0.336; worst q = 0.026; Supplementary Table 8).

Adding the insula to the removed set changed nothing, and removing the entire frontal lobe with cingulate, insular and paracentral cortex (82 regions) left eight of nine system-by-construction tests significant. Within the territory alone, all nine correlations were negative (H3 r = -0.53 to -0.63) with corrected support in one test. Only removal of both poles of the gradient reduced support substantially (five of nine tests). The alignment therefore peaks in frontocingulate cortex and is consistent with a low-frequency, cortex-wide pattern anchored at both ends. The liability maps followed a signed axis between the two poles at Spearman rho = -0.43 to -0.49, within the range expected of maps with their spatial autocorrelation (Moran P = 0.19 to 0.49; Supplementary Table 9).

Residualising target and receptor maps on the nine geometric terms left H3 significant in all nine construction-by-procedure cells (Supplementary Table 10). Its partial correlations were -0.396 to -0.308, with 66% of the raw effect retained. VAChT and NMDA retained 47% and 62% of their effects and four of nine cells each. Across the full family, 27 of 57 tests remained significant under each variogram-matched procedure (26 under both) and 19 of 57 under Moran randomisation. In the positional comparison, the normative child reading-structure map of the CMI cohort correlated with the liability targets at r = -0.221 to -0.236 (eight of nine cells; Supplementary Table 11). The nine geometric terms accounted for 42.7% of that map’s variance and 44.3% of the education-adjusted liability map’s. After the identical adjustment the partial correlations fell to -0.006 to +0.038, with zero cells retained. The Stanford cohort map gave r = -0.270 to -0.352 (2 of nine cells) and partial correlations of -0.074 to -0.132.

As targets, the three reference maps correlated with the liability constructions at |r| = 0.055 to 0.227, with zero of nine cells significant and a best q of 0.461 (Supplementary Table 12). With all three removed simultaneously as covariates, H3 remained significant in every cell (partial r = -0.592 to -0.475, 99% of the raw effect), VAChT in every cell (102%) and NMDA in four of nine (78%). The VAChT value above 100% is a suppression effect, with a larger partial than raw correlation in two of three constructions. The component of H3 and VAChT topography that liability follows therefore lies outside the general molecular topography these maps carry.

The candidate maps are partly correlated (pairwise r = 0.192 to 0.804; H3 with VAChT, 0.615). Removing each map’s leave-one-out common axis alone left two of 57 tests significant under Moran and default variogram-matched procedures, both H3 and VAChT in the unadjusted construction. Under the distribution-matched variant three of 57 remained, the third a positive 5-HT2A partial correlation (r = 0.334, q = 0.0494). After simultaneous adjustment for the common axis and the geometric terms, no receptor was significant in any construction under any procedure (minimum q = 0.0969, 0.0627 and 0.0883). H3 is the only map significant in every cell under the uncontrolled, position-controlled and reference-controlled regimes alike.

The three retained maps are correlated (H3 with VAChT r = 0.615), so partial correlations and Williams tests addressed whether they can be told apart (Supplementary Table 13). In the unadjusted construction, the partial correlation of liability with H3 given VAChT was -0.436 (Moran P = 0.014) and with VAChT given H3 -0.243 (P = 0.111). The Williams test for H3 against VAChT gave t = -1.89 (P = 0.061) at n = 218 and t = -0.77 (P = 0.445) at the effective n of 39. At the nominal parcel count H3 differed from NMDA in every construction and from VAChT only in the education-adjusted construction (unadjusted P = 0.061). No difference held at the effective n.

Sensitivity analyses kept H3 significant under every check, while VAChT and NMDA lost significance under some (Supplementary Tables 14 to 17). VAChT rebuilt without any one of its source studies and restriction to the 210 cortical parcels kept the three systems significant in every construction. Hemisphere-preserving surface spins kept H3 and NMDA significant in every construction and VAChT in two of three constructions. Adult vascular and metabolic covariates left H3 significant in every construction and VAChT and NMDA in two of three constructions. With the nine geometric terms added, H3 remained significant in every construction, NMDA in two of three constructions and VAChT in one of three constructions. Screening 200 pseudo-targets gave per-test rejection rates of 0.043, 0.045 and 0.048 at alpha = 0.05 under the three procedures (Supplementary Table 18). A known driver injected into 1 000 surrogate targets at the observed effect sizes tested recovery (Supplementary Table 19). An H3 driver reached q < 0.05 in 99.8% of draws for the unadjusted construction and in 50 to 55% for the adjusted constructions. It ranked first in 50 to 82% of draws, a VAChT driver in 20 to 47% and an NMDA driver in 2 to 20%. With the shared axis as driver, some map reached q < 0.05 in 74 to 84% of draws, and a retained map ranked first in 8 to 10%. Removing each region’s fractional membership in the 14 resting-state networks of Shirer et al. [26], which explains 27 to 31% of the liability variance, tested a functional confound. H3 and VAChT remained significant in every cell and NMDA in seven of nine (Supplementary Table 20). In the 169 children of ds001894, the two Jacobian definitions gave concordant reading-skill maps (r = 0.916; Supplementary Table 21 and 22). H3 showed the largest prespecified association (r = -0.365; Moran P = 0.039, q = 0.116 across the three prespecified maps and 0.246 across nineteen). NMDA and VAChT correlated at r = - 0.145 and -0.074, and the similarity of the skill map to the three adult liability maps was r = 0.19 to 0.27 (q = 0.55; Supplementary Table 23). A stricter registration threshold of 0.66 (152 children) gave H3 r = -0.316 (Moran P = 0.16; Supplementary Table 24).

In the 9 013 children of the ABCD Study, the smallest corrected probability in the 38-test family was q = 0.909 under Moran randomisation (Supplementary Table 25). Under the variogram-matched procedures it was 0.980 and 0.975. The prespecified correlations were H3 r = -0.172 for adjusted surface area (Moran P = 0.155) and -0.179 for adjusted thickness (P = 0.176). NMDA gave r = 0.009 and -0.023 and VAChT r = 0.183 and - 0.073, and the unadjusted maps gave the same result. Similarity between the skill maps and the adult liability maps was |r| <= 0.21 (best q = 0.84; Supplementary Table 26). Reading skill occupies the lateral temporal and ventral occipitotemporal axis, with the opposite sign in the medial frontal territory where liability peaks.

For the unadjusted construction, a formal comparison of the adult and child H3 correlations gave Williams t = -3.86 (P = 0.0001) at n = 218 (Supplementary Table 27). At the effective n it gave t = -1.58 (P = 0.122). The child map’s regional reliability was 0.48, and disattenuation moved its H3 correlation from -0.365 to -0.527. At the corrected threshold the analysis had power 0.32 for the weakest adult H3 correlation, 0.54 for the strongest and 0.18 for its own effect. Under this power model the child data were underpowered for the adult effect.

A prespecified per-person analysis in the 169 children related each child’s alignment score, the correlation of the child’s regional deviation vector with a receptor map, to reading skill (Supplementary Table 28). The prespecified direction was that a more liability-like anatomy, a more negative score, would go with lower reading skill. The observed association ran the other way. The H3 score predicted the TOWRE score at beta = -3.41 points per SD (95% interval -5.90 to -0.93; HC3 P = 0.0078; permutation P = 0.0098; q = 0.031). The shared-axis score gave beta = -3.10 (q = 0.054), and the VAChT and NMDA scores were near zero. The H3 association kept its direction and size across the sensitivity specifications, and lost corrected significance at the stricter registration threshold (q = 0.088). Children whose volume was relatively lower where H3 density is high therefore read better, the direction of the group-level child skill map and the opposite of the adult liability signature.

The screening of case-control and task-fMRI representations of dyslexia, the source-map uncertainty analysis, the summary-statistics genetic comparison and all sensitivity analyses are reported in the Supplementary Information (Supplementary Methods; Supplementary Tables 29 to 34). A stratified phenotype family of 18 tests gave no q below 0.94. An exploratory triangulation with 22 preselected language regions gave one positive VAChT thickness correlation (r = 0.622, q = 0.035) that failed under rank-based inference (Spearman P = 0.075).

## Discussion

In this study we compared the brain signature of genetic liability for dyslexia with normative maps of neurotransmitter systems. The signature followed the shared geography of the systems that set attention, arousal and synaptic plasticity during learning. Receptor atlases have been related to polygenic risk in Alzheimer’s disease [17] and the genetics of reading to cortical cell types [18]. To our knowledge this is the first study to relate the liability map of a learning disorder to receptor and transporter density. To our knowledge, it is also the first appearance of the histamine H3 receptor in the dyslexia literature, on the searches recorded in the Supplementary Methods.

Each of the three systems that carried the alignment provides one condition of learning. The H3 receptor gates the release of acetylcholine and several other transmitters [36], and postsynaptic H3 receptors on basal-forebrain cholinergic neurons constrain memory in mice [37]. Raising histaminergic tone reshapes learning dynamics in humans [38]. Basal-forebrain acetylcholine sets attention and the readiness of the cortex to change [39, 40], and cholinergic input is required early in learning in mice [41]. Because the alignment belongs to what these maps share, the three systems mark one shared neuromodulatory territory, with no individual system separable from the rest at the effective sample size.

The H3 alignment kept two thirds of its size after the nine geometric terms were removed. It kept almost all of it after blood flow, glucose metabolism and the cognitive component were removed together. It kept about three quarters after membership in 14 resting-state networks was removed. It gave way once the component common to all nineteen maps was removed, and entirely when position was removed with it. The shared component itself aligned with liability in every construction under the variogram-matched procedures. The three retained maps are not its strongest loadings. The liability signature therefore follows an architecture that the neuromodulator systems share [10, 13].

The anatomy of liability appears separate from the anatomy of skill, and where the two meet they carry opposite signs. Reading skill lives in lateral temporal and occipitotemporal tissue [31], while genetic risk leaves its signature along a cingulate and medial frontal, neurotransmitter-rich axis. The watershed and multiple-deficit models describe how many small risks converge on one outcome [42, 43], and they make no spatial claim. We extend them to anatomy. If risk enters through many small upstream paths and difficulty appears downstream where they converge, the skill map should mark the point of convergence. The liability map should mark where the risk enters, in the tissue that prepares the reading circuit to learn. The formal comparison separates them at the nominal but not at the effective sample size (Results), so this extension is ours and stands as a hypothesis. The normative NMDA correspondence with reading skill in children is positive and ventral occipitotemporal [16]. The present NMDA correlation is negative and frontocingulate, and neither of our child cohorts reproduced the NMDA result. The opposite sign is expected where risk lowers volume and skill raises it, so the contrast lies in territory and in replication. The per-person analysis points the same way. Its prespecified direction failed, and the reverse association was significant. Children whose volume is relatively lower where H3 density is high read better, the opposite sign to the adult liability signature. On this reading, molecular involvement appears in the topography of liability before it appears in the physiology of the patient, a sequence these data do not test [44, 45].

The positional comparison sharpens this reading. The normative reading-structure maps of typically developing children relate to the liability targets [16]. That relation dissolved once the nine geometric terms were removed, under the adjustment that left the receptor alignment standing. Liability shares position with the anatomy of reading, and it shares a finer structure with the neuromodulator maps that survives the removal of position.

The frontocingulate territory where liability peaks is the tissue of effortful attention, monitoring and automatisation. Automatisation difficulties are documented as an independent strand in dyslexia [46, 47]. Polygenic scores add predictive information for reading outcomes [48], and the present result suggests why, since they index a layer distinct from the skills that behavioural assessment measures.

The system-level answer yields predictions that anatomy alone leaves unstated. Dyslexia-associated variants should be enriched in genes expressed in histaminergic, cholinergic and glutamatergic signalling, a test that cell-type enrichment methods already applied to reading genetics make feasible [18]. These systems are group-level correlates in adults, and the present data support no pharmacological inference. Whether any of them is a target is a question for mechanistic work with individual-level measurements. The direct test of the present claim is a liability map in children, which recent work shows to be feasible [15]. A per-person alignment score related to reading outcomes would complete that test. Genome-wide studies of dyscalculia and developmental coordination disorder are at an earlier stage, and the first genome-wide study of motor coordination has only recently appeared [49]. Once liability maps for these disorders exist, whether they follow the same axis will be the first transdiagnostic test of this finding.

## Limitations

The alignment belongs to the shared architecture of the nineteen maps, and no individual receptor system can be attributed on this evidence. Partial correlations and Williams tests separate H3 from NMDA at the nominal sample size, and from VAChT in one construction, and no difference holds at the effective sample size. The shared component with position removes every individual result, and the three are not the strongest loadings on that component. The H3 map rests on 8 participants and the VAChT map on three studies of 4, 5 and 18, none with a source-variance image, so no source uncertainty could be propagated. The NMDA map indexes open-channel binding of [18F]GE-179, whose specificity is debated, so it is a putative NMDA-sensitive signal [22, 23]. The effective number of independent regions is about 39. The effective sample size, the reliability estimate and the power model are approximations, so the child comparisons are sensitivity calculations. The three constructions are one map at three depths of adjustment, and the third adjusts for education and verbal-numerical reasoning that may lie downstream of liability. The three randomisation procedures disagree on individual maps, and their empirical size at alpha = 0.05 was 0.043, 0.045 and 0.048. Surface-based spins and adult vascular and metabolic covariates kept H3 in every construction, while VAChT and NMDA lost significance under some (Results), and geodesic-distance surrogates were not applied. The discovery GWAS, the imaging cohort of white British adults aged 45 to 82 and the receptor atlases from younger, largely European donors restrict every result to that population. The child analyses were underpowered for the adult effect and the comparison is a cross-age contrast, so the separation between liability and skill anatomy is descriptive. The per-person association was significant in the direction opposite to the prespecified one and awaits replication.

This study places the brain signature of genetic liability for dyslexia within the neurotransmitter systems that prepare the cortex to learn. It names the three systems in which that architecture is read most clearly, and the shared component itself. Dyslexia research gains a testable biological axis beyond the reading circuit. Inherited risk for dyslexia follows the shared topography of the chemistry of learning, and that correspondence is where the next tests begin.

## Data Availability

This study used only de-identified, publicly available human datasets. Dyslexia-liability maps are available from Dryad (https://doi.org/10.5061/dryad.80gb5mkz6), and the receptor atlas is available from Zenodo (https://doi.org/10.5281/zenodo.20574388). Reference maps were obtained through neuromaps. The child imaging dataset and task-fMRI dataset are available from OpenNeuro as ds001894 and ds003126, respectively. The coordinate archive is available from Dryad (https://doi.org/10.5061/dryad.0p2ngf222). Additional source data are available from the cited publications and public summary-statistics repositories. No new participant data were collected.

https://doi.org/10.5281/zenodo.20574388

https://openneuro.org/datasets/ds001894

https://openneuro.org/datasets/ds003126

https://doi.org/10.5061/dryad.0p2ngf222

## Acknowledgements

This research received no specific grant from any funding agency in the public, commercial, or not-for-profit sectors. The authors thank the investigators who made the reanalysed datasets publicly available: Soheili-Nezhad and colleagues (Dryad), Hansen and colleagues and Zhang and colleagues (Zenodo), the neuromaps developers, the contributors of OpenNeuro datasets ds001894 and ds003126, and the ABCD, GenLang and ENIGMA consortia. Computational resources were provided by the Turkish National Academic Network and Information Center (TUBITAK ULAKBIM) High Performance and Grid Computing Center (TRUBA). Generative AI (Claude, Anthropic) was used with full human oversight for analysis code and language editing; all analytical decisions, interpretations and text are the authors’ own.

## Author contributions

MCC: conceptualisation, methodology, software, formal analysis, data curation, visualisation, writing - original draft, writing - review and editing. YC, MAS, DA and ATA: validation, writing - review and editing. HAV: conceptualisation, methodology, supervision, writing - review and editing. All authors read and approved the final manuscript.

## Ethics statement

This study analysed de-identified, publicly available human data under the access terms of each source repository. No new human material or identifiable participant data were collected. Ethics approval and new informed consent were therefore not applicable to this secondary analysis. The source studies report their own approvals and consent procedures.

## Data availability

Liability maps are available from Dryad (doi:10.5061/dryad.80gb5mkz6) and the receptor atlas from Zenodo (doi:10.5281/zenodo.20574388). Reference maps were obtained through neuromaps, the child cohort from OpenNeuro (ds001894), the task-based fMRI data from OpenNeuro (ds003126), the coordinate archive from Dryad (doi:10.5061/dryad.0p2ngf222), the published child map from the supplement of Carrion-Castillo et al. [31], and the external genetic comparison from the GenLang and ENIGMA3 public summary statistics.

## Code availability

All analysis and verification code, result tables, prespecification documents, seeds and run metadata accompany this submission as the code and data archive and will be deposited at Zenodo before publication, with the DOI inserted at proof. The deposit includes the inputs of the primary screen and the dyslexia_nt package, so the verification script, the territory tests and the null-size, gradient, injection-recovery, per-person and network-membership analyses run from the deposit alone. The figure script, the surface-based analyses and the source-map analyses additionally require the raw atlas files and the neuromaps distribution listed under Data availability.

Supplementary information is available at MP’s website.

