## Supplementary Information for "Mapping the brain signature of dyslexia risk onto histamine, acetylcholine and glutamate receptor systems"

#### Supplementary Methods

**Registration quality in children.** Registration quality in OpenNeuro ds001894 (Lytle et al., 2019) was the correlation between each warped image and the study template. The distribution was bimodal, with 169 scans between 0.633 and 0.721 and 19 scans between -0.150 and 0.525 and none in between, so any threshold between those clusters retains the same 169 children. Sensitivity models re-estimated the reading-skill map with the Woodcock-Johnson III (WJ-III) Broad Reading score in place of the Test of Word Reading Efficiency (TOWRE), with full-scale IQ, with magnetic resonance imaging quality metrics, with quadratic age, with the global covariate omitted and with registration and image-quality outliers removed by a median-absolute-deviation rule over six quality variables. A stricter registration threshold of 0.66, which retains 152 children, was rerun with the primary model on the analysis cluster (Supplementary Table 24).

**Effective sample size, separability and difference tests.** For every primary test, the effective sample size was back-solved as the $n$ at which the two-sided $t$ test of the observed correlation gives the observed surrogate probability (Supplementary Table 3). Dependence among the three retained maps was addressed with partial correlations, each with a probability from 5 000 Moran surrogates of the liability map residualised on the same covariate, and with Williams tests for two dependent correlations sharing the liability map, evaluated at $n = 218$ and at $n = 39$ (Supplementary Table 13). The same Williams test compared the adult liability correlation with H3 against the child reading-skill correlation with H3, with the correlation between the two maps as the dependence term. The reliability of each child map was estimated from its regional model as one minus the mean squared standard error over the variance of the regional estimates, and used to disattenuate the child correlation and to compute power at the corrected threshold of the prespecified family from the Moran null distribution of the child map (Supplementary Table 27).

**Sensitivity analyses added after review.** VAChT was rebuilt leaving out each of its three source studies in turn and from each study alone, with the same participant-number weights (Supplementary Table 14). The prespecified tests were repeated on the 210 cortical parcels with the amygdala and hippocampal subregions removed (Supplementary Table 15). The empirical size of the three randomisation procedures was checked with 200 pseudo-targets, variogram-matched surrogates of the unadjusted liability map that carry its spatial structure and no relation to any receptor map, each screened against the nineteen maps with 500 surrogates under each procedure (Supplementary Table 18). Recovery of a known driver was tested by injecting H3, VAChT, NMDA and the shared axis into Moran surrogates of each construction at the observed correlation, with 1 000 draws per condition. Each pseudo-target was screened against the nineteen maps with 1 000 Moran surrogates and false discovery control across the nineteen tests. The rates at which the driver reached $q < 0.05$, ranked first and ranked within the top three were recorded (Supplementary Table 19).

**Functional-network membership as a covariate.** Each Brainnetome region's fractional overlap with the 14 resting-state networks of Shirer et al. (2012), supplied as MNI152 volumes, was computed at 2 mm. The 14 fractions were removed from the liability constructions and from every receptor map. The partial correlations were tested with 5 000 Moran and 2 000 + 2 000 variogram-matched surrogates of the residualised liability map, with false discovery control across the 57 tests within each procedure (Supplementary Table 20). The fractions explain 27 to 31% of the variance of the three constructions.

**Shared axis, gradient and per-person alignment.** The first principal component of the nineteen standardised maps was oriented so that H3 loads positively and tested as a target under the three procedures (Supplementary Table 5); its loadings are listed in Supplementary Table 6. The gradient description of the Results was tested in two ways (Supplementary Table 9), by the variance of each map explained by the smoothest Moran modes of the region geometry and by the Spearman correlation of each map with a signed axis running from the occipital and inferior temporal pole parcels to the frontocingulate-paracentral territory, with probabilities from 5 000 Moran surrogates of the map. The per-person analysis followed a prespecification written on 6 September 2026 before any score was computed (PRESPECIFICATION_2026-09-06_per_person_alignment.md in the archive). For each of the 169 children, the 218 regional log-Jacobians were residualised across children on age, sex and mean brain log-Jacobian and standardised, and the child's deviation vector was correlated across regions with the H3, VAChT and NMDA maps and the shared axis. Reading score was regressed on each score with the same covariates, with HC3 standard errors and 5 000 subject permutations, and false discovery control across the four scores; the prespecified direction was a positive coefficient (Supplementary Table 28).

**Surface-based nulls and adult physiological covariates.** The Brainnetome volume was carried to the fsaverage surface (10k vertices) by the majority label within a 2 mm ball around each vertex, which gives a surface parcellation of the 210 cortical parcels. An earlier transfer averaged the categorical labels and mislabelled 47% of fsaverage vertices and 39% of fsLR vertices; the corrected transfer changed the decisions for VAChT under the surface spins and for NMDA under the adult covariates (surface_label_transfer_check.json in the archive). Each liability construction was rotated 5 000 times with hemisphere-preserving spins (neuromaps, Alexander-Bloch procedure) and correlated with the nineteen maps, with false discovery control across the 57 tests (Supplementary Table 16). Adult maps of cerebral blood flow, blood volume, oxygen metabolism and glucose metabolism (Raichle and colleagues, distributed by neuromaps on the fsLR 164k surface) were parcellated directly on that surface with the same majority-label transfer of the atlas, and the prespecified H3, VAChT and NMDA tests were repeated with liability and receptor maps residualised on the four adult maps, alone and together with nine geometric terms, with probabilities from 5 000 Moran surrogates of the residualised target on the 210 cortical parcels (Supplementary Table 17).

**Representation specificity.** Two further representations of dyslexia were screened against the nineteen maps in the same parcellation. Case-control maps were reconstructed from the coordinate archive of Yan et al. (2021), which holds 119 experiments and 1 151 peaks. Of these, 92 functional experiments contribute 1 003 peaks and 27 structural experiments contribute 148 peaks. Talairach coordinates were converted to Montreal Neurological Institute (MNI) space (Lancaster et al., 2007). At each region centroid, signed effects were reconstructed with a 20-mm Gaussian kernel. The largest absolute contribution within an experiment was retained, and experiments were combined with effective-sample-size weights. Functional and structural maps were analysed separately. The same coordinates processed with AES-SDM (Radua et al., 2012) gave pooled unthresholded maps that served as a method sensitivity. A participant-level map came from OpenNeuro ds003126 (Banfi et al., 2021), in which 22 typical readers and 20 children with dyslexia read words and pseudohomophones aloud during functional magnetic resonance imaging (fMRI). After motion quality control, 20 typical readers and 13 children with dyslexia formed the group map, adjusted for age, sex and mean framewise displacement. Probabilities for these screens used 5 000 Moran surrogates, with false discovery control across the nineteen maps within each representation. The three liability constructions were then related to the three dyslexia maps under the three procedures, with false discovery control across the nine tests, and to case-control synthesis strata in a stratified family of 18 tests.

**Source-map uncertainty.** Matched group-mean and regional-SD images exist in the atlas source studies for six systems (Hansen et al., 2022). Each source mean was tested against the three constructions under the three procedures, with false discovery control across the 18 tests. In addition, 5 000 perturbations propagated the reported group-mean uncertainty of each source image, and sign stability is the share of perturbations in which the correlation kept its sign. Agreement is the correlation between the source reconstruction and its atlas counterpart.

**External genetic comparison.** Word-reading genome-wide association study (GWAS) statistics (GenLang; Eising et al., 2022) were paired with ENIGMA3-only regional cortical GWAS excluding UK Biobank (Grasby et al., 2020) in cross-trait linkage disequilibrium (LD) score regression (Bulik-Sullivan et al., 2015), with the 1000 Genomes European reference. The regional genetic correlations, multiplied by −1 to carry the dyslexia-liability orientation, formed maps over 34 bilateral Desikan-Killiany regions for surface area and thickness. The nineteen maps were rebuilt at the same regions. All 136 regional models completed with every |*rg*| below 1 and a largest standard error of 0.188. One region, frontal-pole thickness, had single-nucleotide polymorphism heritability *z* of 1.45, and its exclusion left every decision unchanged. An exploratory triangulation related the multivariate GenLang genetic correlations with 22 preselected UK Biobank language-region imaging traits to the three prespecified systems.

**Pipeline verification and prespecification.** Reproduction of the 38 published coefficients of Zhang et al. (2026) had a largest absolute discrepancy of 6.4 x 10^-16, with every published decision matched. The spatial transform of the liability maps was verified against an image available in both native and MNI space ($r = 1.000$ across 1 978 197 voxels). The surface reconstruction recipe reproduced the packaged 68-region Desikan-Killiany receptor matrix at $r \geq 0.997$ for every system; the 74-region Destrieux reconstruction used for the published child maps applies the same recipe and has no independent published reference against which to be checked. The territory analysis, the comparison with dyslexia maps, the stratified family and the two published-map families were prespecified in dated documents that accompany the code, one with a recorded addendum that overrode the parent document's commitment to no third family; the primary screen and the three control families were run first and have no separate document. Every analysis ran with fixed seeds, and the run metadata accompany the code. The Moran spectral randomisation used throughout is a reimplementation of the singleton procedure of BrainSpace; with the same seed it reproduces BrainSpace's surrogates for the three constructions to numerical precision (null standard deviation of *r* identical to seven decimals, identical *P* values for H3, VAChT and NMDA; moran_benchmark_brainspace.csv in the archive). In total the study comprises 26 test families, 745 tests and 1 842 probabilities in 91 false discovery families (Supplementary Table 2).

### Supplementary Results

**Representation specificity.** Across the nineteen maps, the smallest corrected *q* was 0.481 for the functional map, 0.925 for the structural map and 0.150 for the task-based fMRI map. The largest absolute correlation in these screens was 0.312. The three liability constructions related to the same three maps at $r = 0.052$ to 0.149, with a best *q* of 0.634 under any procedure (Supplementary Table 29). The stratified family of 18 tests gave a smallest *q* of 0.94 and a largest absolute correlation of 0.103 (Supplementary Table 34). The receptor alignment therefore belongs to the liability representation of dyslexia.

**Gradient and per-person alignment.** The three liability maps followed the signed pole axis at Spearman $\rho = -0.43$ to -0.49 with Moran $P = 0.19$ to 0.49, so the trend is within the range expected of maps with their spatial autocorrelation; the H3 map followed the same axis at $\rho = 0.71$ ($P = 0.0002$). The smoothest three Moran modes carried 24 to 31% of the variance of the liability maps (Supplementary Table 9). In the per-person analysis (Supplementary Table 28), the prespecified direction was a positive coefficient and the observed coefficients were negative. The H3 score predicted the TOWRE score at $\beta = -3.41$ points per SD (95% interval -5.90 to -0.93; HC3 $P = 0.0078$; permutation $P = 0.0098$; $q = 0.031$) and the shared-axis score at β = -3.10 ($q$ = 0.054); the VAChT and NMDA scores were near zero. The H3 association held across the nonlinear-only definition, WJ-III Broad Reading, quadratic age and the 0.66 registration threshold (β = -3.44 to -2.69; $q$ up to 0.088). Scores were uncorrelated with the child's mean brain log-Jacobian ($|r| \leq 0.02$).

**Source-map uncertainty.** NMDA is the only retained system with a matched source image (Supplementary Table 30); no source uncertainty could be propagated for H3 or VAChT, whose source images carry no variance estimate. Its source mean kept the negative sign in at least 99.5% of perturbations. It reached $q$ = 0.045 in the unadjusted construction under both variogram-matched procedures and $q$ = 0.30 under Moran randomisation.

**External genetic comparison.** Across the 38-test family (Supplementary Table 32), DAT for surface area ($r$ = -0.596) reached $q$ = 0.030 and 0.023 under the two variogram-matched procedures and $q$ = 0.360 under Moran randomisation. In the exploratory 22-region triangulation (Supplementary Table 33), VAChT thickness gave $r$ = 0.622 (Moran $q$ = 0.035 across six tests), held under inverse-variance weighting ($r$ = 0.631) and leave-one-pair omission (0.427 to 0.674), and failed under rank-based inference (Spearman $r$ = 0.470, $P$ = 0.075). This comparison operates at summary-statistics level across 34 bilateral regions and uses reading-ability genetics in place of dyslexia-liability scoring. It tests generalisation across construct, modality and resolution at once.

**Literature search behind the H3 statement.** The Discussion states that the histamine H3 receptor has not previously appeared in the dyslexia literature. The statement rests on searches run on 2026-09-07, recorded with their query strings, record identifiers and titles in literature_search_h3.json in the archive. In PubMed the query ("histamine H3" OR "H3 receptor" OR "H3 receptors" OR H3R OR "histamine 3 receptor") AND (dyslexia OR "reading disability" OR "reading disorder" OR "reading skill" OR "reading ability" OR "word reading") returned 0 records, and the query ("histamine H3" OR "H3 receptor" OR H3R) AND (dyslexia OR dyslexic) returned 0. The broader query (histamine) AND (dyslexia OR dyslexic) returned 1 record, a 1991 case series of antimotion-sickness medications in children with learning disabilities or dyslexia, which does not concern the H3 receptor. In Europe PMC the query ("histamine H3" OR "H3 receptor" OR "H3 receptors" OR H3R) AND (dyslexia OR dyslexic) returned 9 records and its full-text form 7. None is a study of the H3 receptor in dyslexia; the records are reviews of other disorders, a conceptual paper on histamine in attention-deficit hyperactivity disorder and congress abstract books. In the 5 of these records available as full text, no sentence contains both terms. The reading-skill query ("histamine H3" OR "H3 receptor" OR "H3 receptors" OR H3R) AND ("reading disability" OR "reading disorder" OR "reading skill" OR "reading ability" OR "word reading" OR "reading acquisition") returned 5 records, none involving dyslexia or a reading measure.

**Data sources.** Liability maps are available from Dryad (doi:10.5061/dryad.80gb5mkz6; Soheili-Nezhad et al., 2024) and the receptor atlas from Zenodo (doi:10.5281/zenodo.20574388; Zhang et al., 2026). Reference maps were obtained through the neuromaps distribution (Markello et al., 2022), the child cohort from OpenNeuro (ds001894), the published child map from the journal supplement of Carrion-Castillo et al. (2023), the coordinate archive from Dryad (doi:10.5061/dryad.0p2ngf222; Yan et al., 2021) and the task-based fMRI data from OpenNeuro (ds003126). The external genetic comparison used the GenLang and ENIGMA3-only public summary statistics. The accompanying archive holds the three liability targets, the 218-region receptor matrix and the distance matrix of the primary screen, all result tables, the analysis and verification scripts and the run metadata.

### Supplementary Tables

**Supplementary Table 1. Source studies and participant numbers behind the nineteen receptor and transporter maps.** Parsed from the reconstruction script; multi-study systems are combined with participant-number weights as in the source atlas.

| System | Source study (tracer) | Participants per study | Total |
|---|---:|---:|---:|
| 5-HT1A | savli2012 (way100635) | 36 | 36 |
| 5-HT1B | savli2012 (p943); gallezot2010 (p943) | 22 + 65 | 87 |
| 5-HT2A | beliveau2017 (cimbi36) | 29 | 29 |
| 5-HT4 | beliveau2017 (sb207145) | 59 | 59 |
| 5-HT6 | radnakrishnan2018 (gsk215083) | 30 | 30 |
| 5-HTT | beliveau2017 (dasb) | 100 | 100 |
| CB1 | normandin2015 (omar) | 77 | 77 |
| D1 | kaller2017 (sch23390) | 13 | 13 |
| D2 | smith2017 (flb457); sandiego2015 (flb457) | 37 + 55 | 92 |
| DAT | dukart2018 (fpcit) | 174 | 174 |
| GABAA | norgaard2021 (flumazenil) | 16 | 16 |
| H3 | gallezot2017 (gsk189254) | 8 | 8 |
| M1 | naganawa2020 (lsn3172176) | 24 | 24 |
| mGluR5 | rosaneto (abp688); dubois2015 (abp688); smart2019 (abp688) | 22 + 28 + 73 | 123 |
| MOR | kantonen2020 (carfentanil) | 204 | 204 |
| NET | ding2010 (mrb) | 77 | 77 |
| NMDA | galovic2021 (ge179) | 29 | 29 |
| VAChT | tuominen (feobv); bedard2019 (feobv); aghourian2017 (feobv) | 4 + 5 + 18 | 27 |
| α4β2 | hillmer2016 (flubatine) | 30 | 30 |

**Supplementary Table 2. Test families and counts.** Every family evaluated, with the number of tests, the randomisation procedures applied and the number of false discovery families within each procedure.

| Family | Tests | Randomisation procedures | Correction families per procedure |
|---|---:|---:|---:|
| Primary screen, 3 constructions x 19 maps | 57 | 3 | 1 |
| Position control | 57 | 3 | 1 |
| Reference-map covariate control | 57 | 3 | 1 |
| Shared component alone | 57 | 3 | 1 |
| Shared component with position | 57 | 3 | 1 |
| Reference maps as substitute targets | 9 | 3 | 1 |
| Normative reading maps, uncontrolled and position-controlled | 12 | 3 | 2 |
| Territory analyses, 5 analyses x 3 systems x 3 constructions | 45 | 3 | 5 |

| Family | Tests | Randomisation procedures | Correction families per procedure |
|---|---|---|---|
| Case-control coordinate maps, 2 modalities x 19 maps | 38 | 1 | 2 |
| Task-fMRI group map, 19 maps | 19 | 1 | 1 |
| Liability maps x dyslexia maps, 3 x 3 | 9 | 3 | 1 |
| Source-map uncertainty, 3 constructions x 6 systems | 18 | 3 | 1 |
| Reading-skill screen, 169 children, 19 maps x 2 definitions | 38 | 3 | 4 |
| Reading-skill screen, 9 013 children, 19 maps x 2 measures | 38 | 3 | 2 |
| Skill-to-liability similarity, 169 and 9 013 children | 9 | 1 and 3 | 2 |
| External genetic comparison, 19 maps x 2 measures | 38 | 3 | 2 |
| Shared molecular axes as targets, 3 constructions x 3 components | 9 | 3 | 1 |
| Partial correlations among the retained maps, 3 constructions x 6 | 18 | 2 | 0 |
| Williams tests (retained pairs and adult versus child), parametric | 12 | parametric | 0 |
| VAChT source jackknife, 3 constructions x 6 maps | 18 | 2 | 0 |
| Cortex-only prespecified tests, 3 x 3 | 9 | 2 | 0 |
| Gradient pole axis, 7 maps | 7 | 1 | 0 |
| Registration threshold 0.66, 19 maps | 19 | 3 | 2 |
| Per-person alignment, 4 scores x 5 specifications | 20 | permutation | 5 |
| Surface spin test, 3 constructions x 19 maps | 57 | 1 | 1 |
| Adult physiological covariates, 3 x 3 x 2 controls | 18 | 1 | 0 |

**Supplementary Table 3. Effective sample size of the primary tests for the three retained systems.** Effective *n* is the sample size at which the two-sided *t* test of the observed correlation would give the observed surrogate probability. Across the 161 primary tests with probabilities above the surrogate floor, the median was 39 (interquartile range 27 to 59); 9 tests sat at the floor of 1/5 001 and give only a lower bound.

| System | Construction | *r* | Moran *P* | Effective *n* | Variogram *P* | Effective *n* | Distribution *P* | Effective *n* |
|---|---|---|---|---|---|---|---|---|
| H3 | Unadjusted log-Jacobian | −0.618 | 0.0002 | 31 | 0.0002 | 31 | 0.0002 | 31 |
| H3 | Raw Jacobian, head size | −0.475 | 0.0006 | 48 | 0.0042 | 34 | 0.0034 | 36 |
| H3 | Log-Jacobian, head size and education | −0.481 | 0.0006 | 47 | 0.0044 | 33 | 0.0046 | 33 |
| VAChT | Unadjusted log-Jacobian | −0.531 | 0.0002 | 44 | 0.0002 | 44 | 0.0002 | 44 |
| VAChT | Raw Jacobian, head size | −0.401 | 0.0012 | 62 | 0.0004 | 74 | 0.0006 | 70 |
| VAChT | Log-Jacobian, head size and education | −0.358 | 0.0054 | 59 | 0.0026 | 69 | 0.0018 | 74 |
| NMDA | Unadjusted log-Jacobian | −0.329 | 0.0004 | 112 | 0.0002 | 123 | 0.0002 | 123 |
| NMDA | Raw Jacobian, head size | −0.288 | 0.0004 | 147 | 0.0002 | 162 | 0.0006 | 138 |
| NMDA | Log-Jacobian, head size and education | −0.252 | 0.0010 | 167 | 0.0006 | 182 | 0.0006 | 182 |

**Supplementary Table 4. Primary screen of the three liability constructions against all nineteen receptor and transporter maps.** Probabilities come from 5 000 surrogates under each procedure, and $q$ values from false discovery control across the 57 tests within each procedure. Correlations are identical across procedures; only the reference distribution differs.

| System | Construction | $r$ | Moran $P$ | Moran $q$ | Variogram $P$ | Variogram $q$ | Distribution $P$ | Distribution $q$ |
|---|---|---|---|---|---|---|---|---|
| 5-HT1A | Unadjusted log-Jacobian | −0.156 | 0.5225 | 0.7784 | 0.4933 | 0.7210 | 0.4867 | 0.7125 |
| 5-HT1A | Raw Jacobian, head size | −0.015 | 0.9820 | 0.9998 | 0.9448 | 0.9737 | 0.9410 | 0.9666 |
| 5-HT1A | Log-Jacobian, head size and education | 0.013 | 0.9960 | 0.9998 | 0.9566 | 0.9737 | 0.9496 | 0.9666 |
| 5-HT1B | Unadjusted log-Jacobian | −0.102 | 0.8446 | 0.9440 | 0.5937 | 0.7880 | 0.5927 | 0.7833 |
| 5-HT1B | Raw Jacobian, head size | −0.153 | 0.6209 | 0.8426 | 0.3989 | 0.6862 | 0.4023 | 0.6627 |
| 5-HT1B | Log-Jacobian, head size and education | −0.155 | 0.7487 | 0.9425 | 0.3999 | 0.6862 | 0.4093 | 0.6627 |
| 5-HT2A | Unadjusted log-Jacobian | 0.061 | 0.7091 | 0.9399 | 0.6481 | 0.8209 | 0.6393 | 0.8050 |
| 5-HT2A | Raw Jacobian, head size | 0.054 | 0.7802 | 0.9425 | 0.6819 | 0.8269 | 0.6727 | 0.8158 |
| 5-HT2A | Log-Jacobian, head size and education | 0.041 | 0.8102 | 0.9425 | 0.7550 | 0.8662 | 0.7614 | 0.8681 |
| 5-HT4 | Unadjusted log-Jacobian | −0.041 | 0.7952 | 0.9425 | 0.7824 | 0.8745 | 0.7868 | 0.8794 |
| 5-HT4 | Raw Jacobian, head size | 0.030 | 0.8342 | 0.9440 | 0.8456 | 0.9211 | 0.8346 | 0.9039 |
| 5-HT4 | Log-Jacobian, head size and education | 0.047 | 0.7678 | 0.9425 | 0.7497 | 0.8662 | 0.7499 | 0.8681 |
| 5-HT6 | Unadjusted log-Jacobian | −0.224 | 0.1178 | 0.3051 | 0.0832 | 0.2062 | 0.0762 | 0.1971 |
| 5-HT6 | Raw Jacobian, head size | −0.239 | 0.0726 | 0.2069 | 0.0576 | 0.1824 | 0.0538 | 0.1703 |
| 5-HT6 | Log-Jacobian, head size and education | −0.221 | 0.1500 | 0.3562 | 0.0788 | 0.2041 | 0.0830 | 0.1971 |
| 5-HTT | Unadjusted log-Jacobian | −0.287 | 0.1232 | 0.3053 | 0.0668 | 0.1966 | 0.0632 | 0.1812 |
| 5-HTT | Raw Jacobian, head size | −0.125 | 0.5255 | 0.7784 | 0.4265 | 0.6946 | 0.4185 | 0.6627 |
| 5-HTT | Log-Jacobian, head size and education | −0.083 | 0.7465 | 0.9425 | 0.6135 | 0.7947 | 0.6047 | 0.7833 |
| CB1 | Unadjusted log-Jacobian | −0.372 | 0.0014 | 0.0073 | 0.0486 | 0.1731 | 0.0498 | 0.1703 |
| CB1 | Raw Jacobian, head size | −0.256 | 0.0014 | 0.0073 | 0.1942 | 0.3953 | 0.1886 | 0.3839 |
| CB1 | Log-Jacobian, head size and education | −0.239 | 0.0024 | 0.0114 | 0.2589 | 0.5090 | 0.2390 | 0.4697 |
| D1 | Unadjusted log-Jacobian | −0.336 | 0.0344 | 0.1089 | 0.0140 | 0.0614 | 0.0124 | 0.0544 |
| D1 | Raw Jacobian, head size | −0.087 | 0.3705 | 0.6901 | 0.5721 | 0.7880 | 0.5561 | 0.7731 |
| D1 | Log-Jacobian, head size and education | −0.070 | 0.4989 | 0.7784 | 0.6657 | 0.8248 | 0.6497 | 0.8050 |
| D2 | Unadjusted log-Jacobian | −0.171 | 0.4841 | 0.7784 | 0.3935 | 0.6862 | 0.3825 | 0.6627 |
| D2 | Raw Jacobian, head size | −0.037 | 0.9418 | 0.9998 | 0.8564 | 0.9211 | 0.8404 | 0.9039 |
| D2 | Log-Jacobian, head size and education | 0.027 | 0.9908 | 0.9998 | 0.9024 | 0.9526 | 0.8876 | 0.9369 |
| DAT | Unadjusted log-Jacobian | −0.271 | 0.2737 | 0.5573 | 0.1422 | 0.3001 | 0.1282 | 0.2706 |
| DAT | Raw Jacobian, head size | −0.056 | 0.9412 | 0.9998 | 0.7598 | 0.8662 | 0.7542 | 0.8681 |
| DAT | Log-Jacobian, head size and education | −0.003 | 0.9998 | 0.9998 | 0.9876 | 0.9876 | 0.9842 | 0.9842 |
| GABAA | Unadjusted log-Jacobian | −0.126 | 0.5203 | 0.7784 | 0.4389 | 0.6949 | 0.4043 | 0.6627 |
| GABAA | Raw Jacobian, head size | −0.101 | 0.5571 | 0.7784 | 0.5101 | 0.7269 | 0.5085 | 0.7246 |
| GABAA | Log-Jacobian, head size and education | −0.108 | 0.5493 | 0.7784 | 0.4873 | 0.7210 | 0.4875 | 0.7125 |
| H3 | Unadjusted log-Jacobian | −0.618 | 0.0002 | 0.0057 | 0.0002 | 0.0028 | 0.0002 | 0.0038 |

| System | Construction | *r* | Moran *P* | Moran *q* | Variogram *P* | Variogram *q* | Distribution *P* | Distribution *q* |
|---|---|---|---|---|---|---|---|---|
| H3 | Raw Jacobian, head size | −0.475 | 0.0006 | 0.0057 | 0.0042 | 0.0249 | 0.0034 | 0.0215 |
| H3 | Log-Jacobian, head size and education | −0.481 | 0.0006 | 0.0057 | 0.0044 | 0.0249 | 0.0046 | 0.0249 |
| M1 | Unadjusted log-Jacobian | −0.086 | 0.3719 | 0.6901 | 0.4633 | 0.7137 | 0.4615 | 0.7110 |
| M1 | Raw Jacobian, head size | −0.096 | 0.3753 | 0.6901 | 0.4093 | 0.6862 | 0.3933 | 0.6627 |
| M1 | Log-Jacobian, head size and education | −0.062 | 0.5599 | 0.7784 | 0.5945 | 0.7880 | 0.6013 | 0.7833 |
| mGluR5 | Unadjusted log-Jacobian | −0.373 | 0.0294 | 0.0986 | 0.0022 | 0.0179 | 0.0018 | 0.0128 |
| mGluR5 | Raw Jacobian, head size | −0.332 | 0.0238 | 0.0848 | 0.0048 | 0.0249 | 0.0048 | 0.0249 |
| mGluR5 | Log-Jacobian, head size and education | −0.325 | 0.0542 | 0.1626 | 0.0084 | 0.0399 | 0.0080 | 0.0380 |
| MOR | Unadjusted log-Jacobian | −0.503 | 0.0012 | 0.0073 | 0.0186 | 0.0757 | 0.0150 | 0.0611 |
| MOR | Raw Jacobian, head size | −0.345 | 0.0074 | 0.0281 | 0.1102 | 0.2512 | 0.1164 | 0.2551 |
| MOR | Log-Jacobian, head size and education | −0.349 | 0.0074 | 0.0281 | 0.1364 | 0.2990 | 0.1162 | 0.2551 |
| NET | Unadjusted log-Jacobian | −0.228 | 0.5375 | 0.7784 | 0.2783 | 0.5289 | 0.2599 | 0.4939 |
| NET | Raw Jacobian, head size | −0.366 | 0.0968 | 0.2627 | 0.0690 | 0.1966 | 0.0636 | 0.1812 |
| NET | Log-Jacobian, head size and education | −0.355 | 0.2392 | 0.5243 | 0.0768 | 0.2041 | 0.0796 | 0.1971 |
| NMDA | Unadjusted log-Jacobian | −0.329 | 0.0004 | 0.0057 | 0.0002 | 0.0028 | 0.0002 | 0.0038 |
| NMDA | Raw Jacobian, head size | −0.288 | 0.0004 | 0.0057 | 0.0002 | 0.0028 | 0.0006 | 0.0057 |
| NMDA | Log-Jacobian, head size and education | −0.252 | 0.0010 | 0.0073 | 0.0006 | 0.0057 | 0.0006 | 0.0057 |
| VAChT | Unadjusted log-Jacobian | −0.531 | 0.0002 | 0.0057 | 0.0002 | 0.0028 | 0.0002 | 0.0038 |
| VAChT | Raw Jacobian, head size | −0.401 | 0.0012 | 0.0073 | 0.0004 | 0.0046 | 0.0006 | 0.0057 |
| VAChT | Log-Jacobian, head size and education | −0.358 | 0.0054 | 0.0237 | 0.0026 | 0.0185 | 0.0018 | 0.0128 |
| α4β2 | Unadjusted log-Jacobian | −0.337 | 0.2725 | 0.5573 | 0.0876 | 0.2080 | 0.0792 | 0.1971 |
| α4β2 | Raw Jacobian, head size | −0.370 | 0.2214 | 0.5047 | 0.0554 | 0.1824 | 0.0512 | 0.1703 |
| α4β2 | Log-Jacobian, head size and education | −0.383 | 0.3969 | 0.7070 | 0.0464 | 0.1731 | 0.0466 | 0.1703 |

**Supplementary Table 5. Shared molecular axes of the nineteen maps as targets.** First and third principal components of the nineteen maps (38.3% and 11.9% of receptor variance) correlated with the three constructions, with false discovery control across the nine tests within each procedure.

| Component | Construction | *r* | Moran *q* | Variogram *q* | Distribution *q* |
|---|---|---|---|---|---|
| PC1 | Log-Jacobian, head size and education | −0.301 | 0.0702 | 0.0471 | 0.0363 |
| PC1 | Unadjusted log-Jacobian | −0.426 | 0.0378 | 0.0054 | 0.0018 |
| PC1 | Raw Jacobian, head size | −0.324 | 0.0378 | 0.0277 | 0.0238 |
| PC3 | Log-Jacobian, head size and education | −0.312 | 0.0756 | 0.0277 | 0.0238 |
| PC3 | Unadjusted log-Jacobian | −0.326 | 0.0702 | 0.0276 | 0.0238 |
| PC3 | Raw Jacobian, head size | −0.322 | 0.0702 | 0.0276 | 0.0238 |

**Supplementary Table 6. Loadings of the nineteen maps on their first principal component.** The component carries 38.3% of receptor variance and is oriented so that H3 loads positively. The three retained maps are not its strongest loadings (H3 ranks 6, NMDA 9 and VAChT 14 of nineteen).

| System | PC1 loading | Rank by absolute loading |
|---|---:|---:|
| mGluR5 | 0.342 | 1 |
| 5-HT6 | 0.310 | 2 |
| CB1 | 0.280 | 3 |
| M1 | 0.279 | 4 |
| D1 | 0.269 | 5 |
| H3 | 0.266 | 6 |
| MOR | 0.253 | 7 |
| GABAA | 0.241 | 8 |
| NMDA | 0.231 | 9 |
| α4β2 | 0.227 | 10 |
| 5-HT1B | 0.219 | 11 |
| 5-HT4 | 0.204 | 12 |
| 5-HT2A | 0.202 | 13 |
| VAChT | 0.191 | 14 |
| D2 | 0.190 | 15 |
| NET | 0.160 | 16 |
| DAT | 0.125 | 17 |
| 5-HT1A | 0.109 | 18 |
| 5-HTT | 0.067 | 19 |

**Supplementary Table 7. Mean standardised H3 density and liability effect by gyral group.** Groups follow Brainnetome index ranges; prefrontal cortex combines the superior, middle, inferior frontal and orbital gyri, and occipital cortex the medioventral and lateral occipital parcels.

| Gyral group | Regions | H3 density (z) | Liability, unadjusted (z) | Liability, head size (z) | Liability, education (z) |
|---|---:|---:|---:|---:|---:|
| Cingulate cortex | 14 | 1.13 | −0.83 | −0.28 | −0.53 |
| Prefrontal cortex | 64 | 0.75 | −0.41 | −0.22 | −0.23 |
| Paracentral lobule | 4 | 0.39 | −1.58 | −2.09 | −2.04 |
| Precentral gyrus | 12 | 0.04 | −0.10 | −0.52 | −0.54 |
| Postcentral gyrus | 8 | −0.20 | 0.01 | −0.35 | −0.31 |
| Superior temporal gyrus | 16 | −0.26 | −0.17 | −0.34 | −0.24 |
| Posterior cingulate, precuneus | 18 | −0.40 | 0.55 | 0.42 | 0.42 |
| Inferior parietal lobe | 12 | −0.45 | 0.12 | −0.23 | −0.14 |
| Middle temporal gyrus | 28 | −0.52 | 0.25 | 0.39 | 0.42 |
| Superior parietal lobule | 10 | −0.64 | 0.18 | −0.32 | −0.42 |

| Gyral group | Regions | H3 density (z) | Liability, unadjusted (z) | Liability, head size (z) | Liability, education (z) |
|---|---|---|---|---|---|
| Inferior temporal gyrus | 20 | −0.74 | 0.65 | 0.73 | 0.82 |
| Occipital gyrus | 12 | −0.98 | 1.27 | 1.16 | 1.11 |

**Supplementary Table 8. Territory analyses.** A construction counts as retained when its test reaches $q < 0.05$ under all three procedures, with false discovery control across the nine prespecified tests of each analysis.

| Analysis | System | *r*, unadjusted | *r*, head size | *r*, education | Constructions retained |
|---|---|---|---|---|---|
| Territory removed (186 regions) | H3 | −0.525 | −0.371 | −0.336 | 3 of 3 |
| Territory removed (186 regions) | NMDA | −0.330 | −0.285 | −0.243 | 3 of 3 |
| Territory removed (186 regions) | VAChT | −0.537 | −0.380 | −0.323 | 3 of 3 |
| Territory and insula removed (174 regions) | H3 | −0.491 | −0.382 | −0.360 | 3 of 3 |
| Territory and insula removed (174 regions) | NMDA | −0.258 | −0.263 | −0.232 | 3 of 3 |
| Territory and insula removed (174 regions) | VAChT | −0.502 | −0.423 | −0.377 | 3 of 3 |
| Frontal lobe, cingulate, insula and paracentral lobule removed (136 regions) | H3 | −0.480 | −0.410 | −0.400 | 3 of 3 |
| Frontal lobe, cingulate, insula and paracentral lobule removed (136 regions) | NMDA | −0.213 | −0.211 | −0.178 | 2 of 3 |
| Frontal lobe, cingulate, insula and paracentral lobule removed (136 regions) | VAChT | −0.540 | −0.444 | −0.400 | 3 of 3 |
| Territory only (32 regions) | H3 | −0.629 | −0.526 | −0.555 | 1 of 3 |
| Territory only (32 regions) | NMDA | −0.526 | −0.416 | −0.442 | 0 of 3 |
| Territory only (32 regions) | VAChT | −0.458 | −0.406 | −0.396 | 0 of 3 |
| Both gradient poles removed (150 regions) | H3 | −0.446 | −0.233 | −0.182 | 1 of 3 |
| Both gradient poles removed (150 regions) | NMDA | −0.380 | −0.288 | −0.245 | 3 of 3 |
| Both gradient poles removed (150 regions) | VAChT | −0.471 | −0.239 | −0.156 | 1 of 3 |

**Supplementary Table 9. Formal description of the cortex-wide gradient.** Moran modes are the eigenvectors of the double-centred inverse-distance weight matrix used by the Moran randomisation, smoothest first. The pole axis is, for every region, the mean distance to the occipital and inferior temporal pole parcels minus the mean distance to the frontocingulate-paracentral territory parcels (positive towards the frontocingulate pole). The Moran *P* refers to the Spearman correlation with that axis under 5 000 Moran surrogates of the map; isotonic $R^2$ is the variance explained by a monotone fit on the axis.

| Map | $R^2$ on Moran mode 1 | $R^2$ on modes 1 to 3 | $R^2$ on modes 1 to 10 | Spearman ρ with pole axis | Moran *P* | Isotonic $R^2$ |
|---|---|---|---|---|---|---|
| Liability, unadjusted | 0.001 | 0.244 | 0.319 | −0.486 | 0.1914 | 0.370 |
| Liability, head size | 0.002 | 0.242 | 0.347 | −0.434 | 0.4109 | 0.326 |
| Liability, education | 0.002 | 0.310 | 0.403 | −0.485 | 0.4943 | 0.363 |
| H3 | 0.034 | 0.430 | 0.444 | 0.709 | 0.0002 | 0.460 |
| VAChT | 0.000 | 0.112 | 0.299 | 0.455 | 0.0002 | 0.245 |
| NMDA | 0.006 | 0.012 | 0.105 | 0.048 | 0.8108 | 0.052 |

| Map | $R^2$ on Moran mode 1 | $R^2$ on modes 1 to 3 | $R^2$ on modes 1 to 10 | Spearman ρ with pole axis | Moran *P* | Isotonic $R^2$ |
|---|---|---|---|---|---|---|
| shared axis | 0.006 | 0.179 | 0.381 | 0.356 | 0.3597 | 0.195 |

**Supplementary Table 10. Control regimes for the three retained systems.** Retention is the mean absolute partial correlation across the three constructions as a percentage of the mean absolute uncontrolled correlation. Cells are construction-by-procedure cells reaching $q < 0.05$, of nine. The VAChT retention above 100% is a suppression effect, in which two of three constructions give a larger partial than raw correlation after the reference maps are removed.

| System | Position control, partial *r* | Retained | Cells | Reference maps removed, partial *r* | Retained | Cells | Shared component alone, cells | Shared component with position, cells |
|---|---|---|---|---|---|---|---|---|
| H3 | −0.396 to −0.308 | 66% | 9 | −0.592 to −0.475 | 99% | 9 | 3 | 0 |
| VAChT | −0.289 to −0.119 | 47% | 4 | −0.518 to −0.378 | 102% | 9 | 3 | 0 |
| NMDA | −0.195 to −0.147 | 62% | 4 | −0.276 to −0.175 | 78% | 4 | 0 | 0 |

**Supplementary Table 11. Normative child reading-structure maps in the positional comparison.** Both cohort maps of Zhang et al. (2026) are shown uncontrolled and after residualisation on the nine geometric terms, with false discovery control across the six tests.

| Map | Construction | *r* | Moran *q* | Variogram *q* | Distribution *q* | Partial *r* after position | Moran *q* | Variogram *q* | Distribution *q* |
|---|---|---|---|---|---|---|---|---|---|
| Normative reading map, CMI sample | Unadjusted log-Jacobian | −0.221 | 0.0948 | 0.0378 | 0.0348 | 0.038 | 0.9734 | 0.9708 | 0.9746 |
| Normative reading map, CMI sample | Raw Jacobian, head size | −0.236 | 0.0210 | 0.0378 | 0.0348 | −0.006 | 0.9734 | 0.9708 | 0.9746 |
| Normative reading map, CMI sample | Log-Jacobian, head size and education | −0.231 | 0.0210 | 0.0378 | 0.0348 | −0.003 | 0.9734 | 0.9708 | 0.9746 |
| Normative reading map, Stanford sample | Unadjusted log-Jacobian | −0.352 | 0.0892 | 0.0378 | 0.0348 | −0.132 | 0.9734 | 0.9708 | 0.9746 |
| Normative reading map, Stanford sample | Raw Jacobian, head size | −0.275 | 0.0948 | 0.0619 | 0.0562 | −0.079 | 0.9734 | 0.9708 | 0.9746 |
| Normative reading map, Stanford sample | Log-Jacobian, head size and education | −0.270 | 0.1116 | 0.0680 | 0.0562 | −0.074 | 0.9734 | 0.9708 | 0.9746 |

**Supplementary Table 12. Reference maps as substitute targets.** The three reference maps served as targets for the three liability constructions, with false discovery control across the nine tests.

| Reference map | Construction | *r* | Moran *P* | Moran *q* | Variogram *q* | Distribution *q* |
|---|---|---|---|---|---|---|
| Neurosynth cognitive component | Unadjusted log-Jacobian | 0.055 | 0.8002 | 0.8002 | 0.7912 | 0.7912 |
| Neurosynth cognitive component | Raw Jacobian, head size | −0.167 | 0.5431 | 0.7174 | 0.4663 | 0.4607 |
| Neurosynth cognitive component | Log-Jacobian, head size and education | −0.170 | 0.6377 | 0.7174 | 0.4663 | 0.4607 |
| Cerebral blood flow | Unadjusted log-Jacobian | −0.186 | 0.3823 | 0.7174 | 0.4663 | 0.4607 |
| Cerebral blood flow | Raw Jacobian, head size | −0.163 | 0.4895 | 0.7174 | 0.4663 | 0.4607 |
| Cerebral blood flow | Log-Jacobian, head size and education | −0.157 | 0.5681 | 0.7174 | 0.4663 | 0.4607 |

| Reference map | Construction | r | Moran P | Moran q | Variogram q | Distribution q |
|---|---|---|---|---|---|---|
| Glucose metabolism | Unadjusted log-Jacobian | −0.193 | 0.4831 | 0.7174 | 0.4663 | 0.4607 |
| Glucose metabolism | Raw Jacobian, head size | −0.218 | 0.4607 | 0.7174 | 0.4663 | 0.4607 |
| Glucose metabolism | Log-Jacobian, head size and education | −0.227 | 0.5225 | 0.7174 | 0.4663 | 0.4607 |

**Supplementary Table 13. Partial correlations and Williams tests among the three retained maps.** Partial correlations control for the other map of the pair; their probabilities use 5 000 Moran surrogates of the liability map residualised in the same way. Williams tests compare the two dependent correlations at the nominal *n* of 218 and at the effective *n* of 39.

| Construction | Pair (A vs B) | r A | r B | r A,B | partial r A given B | Moran P | partial r B given A | Moran P | Williams t (n = 218) | P | Williams t (n = 39) | P |
|---|---|---|---|---|---|---|---|---|---|---|---|---|
| Unadjusted log-Jacobian | H3 vs VAChT | −0.618 | −0.531 | 0.615 | −0.436 | 0.0138 | −0.243 | 0.1106 | -1.89 | 0.0607 | -0.77 | 0.4451 |
| Unadjusted log-Jacobian | H3 vs NMDA | −0.618 | −0.329 | 0.346 | −0.569 | 0.0002 | −0.156 | 0.1368 | -4.63 | <0.0001 | -1.90 | 0.0658 |
| Unadjusted log-Jacobian | VAChT vs NMDA | −0.531 | −0.329 | 0.455 | −0.453 | 0.0010 | −0.117 | 0.2615 | -3.32 | 0.0011 | -1.36 | 0.1831 |
| Raw Jacobian, head size | H3 vs VAChT | −0.475 | −0.401 | 0.615 | −0.316 | 0.3803 | −0.157 | 0.4865 | -1.41 | 0.1590 | -0.58 | 0.5665 |
| Raw Jacobian, head size | H3 vs NMDA | −0.475 | −0.288 | 0.346 | −0.418 | 0.0038 | −0.150 | 0.0692 | -2.71 | 0.0072 | -1.11 | 0.2737 |
| Raw Jacobian, head size | VAChT vs NMDA | −0.401 | −0.288 | 0.455 | −0.317 | 0.0312 | −0.130 | 0.1814 | -1.73 | 0.0843 | -0.71 | 0.4823 |
| Log-Jacobian, head size and education | H3 vs VAChT | −0.481 | −0.358 | 0.615 | −0.355 | 0.3471 | −0.090 | 0.5989 | -2.35 | 0.0198 | -0.96 | 0.3432 |
| Log-Jacobian, head size and education | H3 vs NMDA | −0.481 | −0.252 | 0.346 | −0.434 | 0.0052 | −0.104 | 0.2364 | -3.32 | 0.0011 | -1.36 | 0.1824 |
| Log-Jacobian, head size and education | VAChT vs NMDA | −0.358 | −0.252 | 0.455 | −0.282 | 0.0582 | −0.108 | 0.2356 | -1.59 | 0.1135 | -0.65 | 0.5196 |

**Supplementary Table 14. VAChT rebuilt leaving out each source study in turn, and each source study alone.** Probabilities from 5 000 Moran and 2 000 variogram-matched surrogates of the liability construction.

| Construction | VAChT map | r | Moran P | Variogram P | r with atlas VAChT |
|---|---|---|---|---|---|
| Unadjusted log-Jacobian | all three (atlas) | −0.531 | 0.0004 | 0.0005 | 1.000 |
| Unadjusted log-Jacobian | without tuominen | −0.527 | 0.0006 | 0.0005 | 0.997 |
| Unadjusted log-Jacobian | without bedard2019 | −0.543 | 0.0004 | 0.0005 | 0.996 |
| Unadjusted log-Jacobian | without aghourian2017 | −0.475 | 0.0006 | 0.0005 | 0.956 |
| Unadjusted log-Jacobian | tuominen only (n = 4) | −0.506 | 0.0002 | 0.0005 | 0.935 |
| Unadjusted log-Jacobian | bedard2019 only (n = 5) | −0.430 | 0.0016 | 0.0005 | 0.931 |
| Unadjusted log-Jacobian | aghourian2017 only (n = 18) | −0.540 | 0.0006 | 0.0005 | 0.988 |
| Raw Jacobian, head size | all three (atlas) | −0.401 | 0.0018 | 0.0005 | 1.000 |
| Raw Jacobian, head size | without tuominen | −0.408 | 0.0020 | 0.0010 | 0.997 |

| Construction | VAChT map | *r* | Moran *P* | Variogram *P* | *r* with atlas VAChT |
|---|---|---|---|---|---|
| Raw Jacobian, head size | without bedard2019 | −0.427 | 0.0018 | 0.0005 | 0.996 |
| Raw Jacobian, head size | without aghourian2017 | −0.304 | 0.0070 | 0.0020 | 0.956 |
| Raw Jacobian, head size | tuominen only (n = 4) | −0.337 | 0.0036 | 0.0005 | 0.935 |
| Raw Jacobian, head size | bedard2019 only (n = 5) | −0.264 | 0.0340 | 0.0260 | 0.931 |
| Raw Jacobian, head size | aghourian2017 only (n = 18) | −0.437 | 0.0018 | 0.0005 | 0.988 |
| Log-Jacobian, head size and education | all three (atlas) | −0.358 | 0.0050 | 0.0030 | 1.000 |
| Log-Jacobian, head size and education | without tuominen | −0.365 | 0.0054 | 0.0030 | 0.997 |
| Log-Jacobian, head size and education | without bedard2019 | −0.387 | 0.0052 | 0.0015 | 0.996 |
| Log-Jacobian, head size and education | without aghourian2017 | −0.254 | 0.0182 | 0.0215 | 0.956 |
| Log-Jacobian, head size and education | tuominen only (n = 4) | −0.298 | 0.0074 | 0.0025 | 0.935 |
| Log-Jacobian, head size and education | bedard2019 only (n = 5) | −0.208 | 0.1116 | 0.0925 | 0.931 |
| Log-Jacobian, head size and education | aghourian2017 only (n = 18) | −0.398 | 0.0056 | 0.0015 | 0.988 |

**Supplementary Table 15. Prespecified tests on the 210 cortical parcels alone.** Amygdala and hippocampal subregions removed; distance matrix restricted accordingly.

| Construction | System | *r* (210 cortical regions) | Moran *P* | Variogram *P* |
|---|---|---|---|---|
| Unadjusted log-Jacobian | H3 | −0.628 | 0.0002 | 0.0005 |
| Unadjusted log-Jacobian | VAChT | −0.531 | 0.0008 | 0.0005 |
| Unadjusted log-Jacobian | NMDA | −0.336 | 0.0014 | 0.0005 |
| Raw Jacobian, head size | H3 | −0.484 | 0.0010 | 0.0100 |
| Raw Jacobian, head size | VAChT | −0.416 | 0.0038 | 0.0020 |
| Raw Jacobian, head size | NMDA | −0.291 | 0.0002 | 0.0005 |
| Log-Jacobian, head size and education | H3 | −0.496 | 0.0010 | 0.0100 |
| Log-Jacobian, head size and education | VAChT | −0.386 | 0.0104 | 0.0070 |
| Log-Jacobian, head size and education | NMDA | −0.252 | 0.0008 | 0.0015 |

**Supplementary Table 16. Surface-based spin test of the primary screen on the 210 cortical parcels.** Parcel values on fsaverage (10k) with 5 000 hemisphere-preserving rotations of each liability construction (Alexander-Bloch procedure, neuromaps); false discovery control across the 57 tests.

| System | Construction | *r* (210 cortical regions) | Spin *P* | Spin *q* (57) |
|---|---|---|---|---|
| 5-HT1A | Unadjusted log-Jacobian | −0.159 | 0.5591 | 0.8171 |
| 5-HT1A | Raw Jacobian, head size | −0.025 | 0.9260 | 0.9763 |
| 5-HT1A | Log-Jacobian, head size and education | −0.002 | 0.9944 | 0.9944 |
| 5-HT1B | Unadjusted log-Jacobian | −0.130 | 0.5523 | 0.8171 |
| 5-HT1B | Raw Jacobian, head size | −0.161 | 0.4183 | 0.7013 |
| 5-HT1B | Log-Jacobian, head size and education | −0.155 | 0.4687 | 0.7633 |

| System | Construction | *r* (210 cortical regions) | Spin *P* | Spin *q* (57) |
|---|---|---|---|---|
| 5-HT2A | Unadjusted log-Jacobian | −0.016 | 0.9320 | 0.9763 |
| 5-HT2A | Raw Jacobian, head size | 0.033 | 0.8430 | 0.9763 |
| 5-HT2A | Log-Jacobian, head size and education | 0.042 | 0.8100 | 0.9763 |
| 5-HT4 | Unadjusted log-Jacobian | −0.072 | 0.7586 | 0.9668 |
| 5-HT4 | Raw Jacobian, head size | 0.023 | 0.9146 | 0.9763 |
| 5-HT4 | Log-Jacobian, head size and education | 0.050 | 0.8352 | 0.9763 |
| 5-HT6 | Unadjusted log-Jacobian | −0.268 | 0.0612 | 0.1938 |
| 5-HT6 | Raw Jacobian, head size | −0.254 | 0.0684 | 0.2052 |
| 5-HT6 | Log-Jacobian, head size and education | −0.223 | 0.1168 | 0.2773 |
| 5-HTT | Unadjusted log-Jacobian | −0.247 | 0.2050 | 0.4327 |
| 5-HTT | Raw Jacobian, head size | −0.095 | 0.6401 | 0.8899 |
| 5-HTT | Log-Jacobian, head size and education | −0.064 | 0.7618 | 0.9668 |
| CB1 | Unadjusted log-Jacobian | −0.364 | 0.0380 | 0.1444 |
| CB1 | Raw Jacobian, head size | −0.247 | 0.2404 | 0.4893 |
| CB1 | Log-Jacobian, head size and education | −0.233 | 0.3447 | 0.6127 |
| D1 | Unadjusted log-Jacobian | −0.323 | 0.0560 | 0.1938 |
| D1 | Raw Jacobian, head size | −0.066 | 0.7632 | 0.9668 |
| D1 | Log-Jacobian, head size and education | −0.052 | 0.8318 | 0.9763 |
| D2 | Unadjusted log-Jacobian | −0.126 | 0.6323 | 0.8899 |
| D2 | Raw Jacobian, head size | −0.005 | 0.9864 | 0.9944 |
| D2 | Log-Jacobian, head size and education | 0.050 | 0.8654 | 0.9763 |
| DAT | Unadjusted log-Jacobian | −0.230 | 0.3281 | 0.6127 |
| DAT | Raw Jacobian, head size | −0.022 | 0.9292 | 0.9763 |
| DAT | Log-Jacobian, head size and education | 0.019 | 0.9420 | 0.9763 |
| GABAA | Unadjusted log-Jacobian | −0.184 | 0.1986 | 0.4327 |
| GABAA | Raw Jacobian, head size | −0.122 | 0.3521 | 0.6127 |
| GABAA | Log-Jacobian, head size and education | −0.114 | 0.3547 | 0.6127 |
| H3 | Unadjusted log-Jacobian | −0.628 | 0.0004 | 0.0076 |
| H3 | Raw Jacobian, head size | −0.484 | 0.0008 | 0.0091 |
| H3 | Log-Jacobian, head size and education | −0.496 | 0.0008 | 0.0091 |
| M1 | Unadjusted log-Jacobian | −0.092 | 0.5243 | 0.8077 |
| M1 | Raw Jacobian, head size | −0.086 | 0.5097 | 0.8070 |
| M1 | Log-Jacobian, head size and education | −0.046 | 0.7265 | 0.9668 |
| mGluR5 | Unadjusted log-Jacobian | −0.431 | 0.0014 | 0.0133 |
| mGluR5 | Raw Jacobian, head size | −0.363 | 0.0074 | 0.0383 |
| mGluR5 | Log-Jacobian, head size and education | −0.345 | 0.0170 | 0.0745 |
| MOR | Unadjusted log-Jacobian | −0.501 | 0.0040 | 0.0285 |
| MOR | Raw Jacobian, head size | −0.345 | 0.1040 | 0.2577 |
| MOR | Log-Jacobian, head size and education | −0.359 | 0.1350 | 0.3077 |

| System | Construction | *r* (210 cortical regions) | Spin *P* | Spin *q* (57) |
|---|---|---|---|---|
| NET | Unadjusted log-Jacobian | −0.258 | 0.2587 | 0.5086 |
| NET | Raw Jacobian, head size | −0.381 | 0.0318 | 0.1294 |
| NET | Log-Jacobian, head size and education | −0.362 | 0.0918 | 0.2491 |
| NMDA | Unadjusted log-Jacobian | −0.336 | 0.0002 | 0.0057 |
| NMDA | Raw Jacobian, head size | −0.291 | 0.0022 | 0.0179 |
| NMDA | Log-Jacobian, head size and education | −0.252 | 0.0052 | 0.0308 |
| VAChT | Unadjusted log-Jacobian | −0.531 | 0.0002 | 0.0057 |
| VAChT | Raw Jacobian, head size | −0.416 | 0.0054 | 0.0308 |
| VAChT | Log-Jacobian, head size and education | −0.386 | 0.0148 | 0.0703 |
| α4β2 | Unadjusted log-Jacobian | −0.360 | 0.1014 | 0.2577 |
| α4β2 | Raw Jacobian, head size | −0.379 | 0.0602 | 0.1938 |
| α4β2 | Log-Jacobian, head size and education | −0.385 | 0.0776 | 0.2211 |

**Supplementary Table 17. Prespecified tests after removal of adult vascular and metabolic maps.** Cerebral blood flow, blood volume, oxygen metabolism and glucose metabolism from the adult PET maps of Raichle and colleagues (neuromaps), parcellated on the surface to the 210 cortical parcels, removed from liability and receptor maps alone and together with nine geometric terms; probabilities from 5 000 Moran surrogates of the residualised target.

| Construction | Covariates removed | System | *r* (210 regions) | Partial *r* | Moran *P* |
|---|---|---|---|---|---|
| Unadjusted log-Jacobian | adult CBF, CBV, CMRO2, CMRglc | H3 | −0.628 | −0.428 | 0.0002 |
| Unadjusted log-Jacobian | adult CBF, CBV, CMRO2, CMRglc | VAChT | −0.531 | −0.328 | 0.0008 |
| Unadjusted log-Jacobian | adult CBF, CBV, CMRO2, CMRglc | NMDA | −0.336 | −0.278 | 0.0056 |
| Unadjusted log-Jacobian | adult physiology and nine geometric terms | H3 | −0.628 | −0.422 | 0.0002 |
| Unadjusted log-Jacobian | adult physiology and nine geometric terms | VAChT | −0.531 | −0.232 | 0.0052 |
| Unadjusted log-Jacobian | adult physiology and nine geometric terms | NMDA | −0.336 | −0.244 | 0.0118 |
| Raw Jacobian, head size | adult CBF, CBV, CMRO2, CMRglc | H3 | −0.484 | −0.307 | 0.0024 |
| Raw Jacobian, head size | adult CBF, CBV, CMRO2, CMRglc | VAChT | −0.416 | −0.249 | 0.0168 |
| Raw Jacobian, head size | adult CBF, CBV, CMRO2, CMRglc | NMDA | −0.291 | −0.205 | 0.0226 |
| Raw Jacobian, head size | adult physiology and nine geometric terms | H3 | −0.484 | −0.340 | 0.0008 |
| Raw Jacobian, head size | adult physiology and nine geometric terms | VAChT | −0.416 | −0.094 | 0.2565 |
| Raw Jacobian, head size | adult physiology and nine geometric terms | NMDA | −0.291 | −0.204 | 0.0300 |
| Log-Jacobian, head size and education | adult CBF, CBV, CMRO2, CMRglc | H3 | −0.496 | −0.299 | 0.0022 |
| Log-Jacobian, head size and education | adult CBF, CBV, CMRO2, CMRglc | VAChT | −0.386 | −0.200 | 0.0610 |
| Log-Jacobian, head size and education | adult CBF, CBV, CMRO2, CMRglc | NMDA | −0.252 | −0.165 | 0.0600 |
| Log-Jacobian, head size and education | adult physiology and nine geometric terms | H3 | −0.496 | −0.328 | 0.0012 |
| Log-Jacobian, head size and education | adult physiology and nine geometric terms | VAChT | −0.386 | −0.024 | 0.7740 |
| Log-Jacobian, head size and education | adult physiology and nine geometric terms | NMDA | −0.252 | −0.177 | 0.0504 |

**Supplementary Table 18. Empirical size of the three randomisation procedures.** Pseudo-targets are variogram-matched surrogates of the unadjusted liability map, each screened against the nineteen maps with 500 surrogates under each procedure.

| **Procedure** | **Pseudo-targets** | **Per-test rejection rate at α = 0.05** | **Share with any map at *q* < 0.05 (19-map family)** |
|---|---|---|---|
| Moran | 200 | 0.043 | 0.035 |
| Variogram-matched | 200 | 0.045 | 0.035 |
| Distribution-matched | 200 | 0.048 | 0.020 |

**Supplementary Table 19. Recovery of a known driver injected into surrogate targets.** Each driver was injected into Moran surrogates of the named construction at its observed correlation, with 1 000 draws per condition. Each pseudo-target was screened against the nineteen maps with 1 000 Moran surrogates and false discovery control across the nineteen tests. For the shared axis, reaching *q* < 0.05 means that any map did so, and ranked first means that H3, VAChT or NMDA had the largest absolute correlation.

| **Driver** | **Construction** | **Injected *r*** | **Realised *r* (mean ± SD)** | **Reached *q* < 0.05** | **Ranked first** | **Within top three** | **Maps most often first** |
|---|---|---|---|---|---|---|---|
| H3 | Unadjusted log-Jacobian | −0.618 | −0.601 ± 0.127 | 0.998 | 0.820 | 0.984 | H3 0.82; D1 0.08; D2 0.03; DAT 0.02 |
| H3 | Raw Jacobian, head size | −0.475 | −0.465 ± 0.175 | 0.550 | 0.525 | 0.641 | H3 0.53; DAT 0.18; D1 0.17; D2 0.10 |
| H3 | Log-Jacobian, head size and education | −0.481 | −0.461 ± 0.195 | 0.502 | 0.496 | 0.532 | H3 0.50; DAT 0.25; D1 0.14; D2 0.10 |
| VAChT | Unadjusted log-Jacobian | −0.531 | −0.523 ± 0.097 | 0.898 | 0.469 | 0.901 | VAChT 0.47; DAT 0.32; H3 0.09; D2 0.04 |
| VAChT | Raw Jacobian, head size | −0.401 | −0.390 ± 0.121 | 0.572 | 0.241 | 0.487 | DAT 0.41; VAChT 0.24; H3 0.21; α4β2 0.08 |
| VAChT | Log-Jacobian, head size and education | −0.358 | −0.352 ± 0.124 | 0.405 | 0.205 | 0.331 | DAT 0.45; VAChT 0.20; H3 0.16; α4β2 0.10 |
| NMDA | Unadjusted log-Jacobian | −0.329 | −0.325 ± 0.094 | 0.594 | 0.197 | 0.504 | NMDA 0.20; α4β2 0.17; DAT 0.13; H3 0.11 |
| NMDA | Raw Jacobian, head size | −0.288 | −0.290 ± 0.072 | 0.718 | 0.087 | 0.360 | α4β2 0.26; DAT 0.20; 5-HT1B 0.14; NET 0.11 |
| NMDA | Log-Jacobian, head size and education | −0.252 | −0.251 ± 0.074 | 0.533 | 0.018 | 0.117 | α4β2 0.34; DAT 0.21; 5-HT1B 0.12; NET 0.12 |
| Shared axis (PC1) | Unadjusted log-Jacobian | −0.426 | −0.419 ± 0.134 | 0.843 | 0.101 | 0.303 | D2 0.20; mGluR5 0.13; D1 0.10; H3 0.10 |
| Shared axis (PC1) | Raw Jacobian, head size | −0.324 | −0.318 ± 0.131 | 0.782 | 0.089 | 0.292 | D2 0.23; α4β2 0.17; 5-HT1B 0.15; D1 0.12 |
| Shared axis (PC1) | Log-Jacobian, head size and education | −0.301 | −0.294 ± 0.148 | 0.743 | 0.079 | 0.339 | D2 0.26; α4β2 0.22; 5-HT1B 0.13; D1 0.10 |

**Supplementary Table 20. Functional-network membership as a covariate.** Each region's fractional overlap with the 14 resting-state networks of Shirer et al. (2012) was removed from the liability constructions and from every receptor map; the fractions explain 30% of the variance of the education-adjusted liability map. Probabilities come from 5 000 Moran and 2 000 + 2 000 variogram-matched surrogates of the residualised liability map, with false discovery control across the 57 tests within each procedure. Cells count construction-by-procedure cells with *q* < 0.05 and a negative partial correlation.

| **System** | **Partial *r* (three constructions)** | **Moran cells (of 3)** | **Variogram cells (of 3)** | **Distribution cells (of 3)** | **Cells of nine** |
|---|---|---|---|---|---|
| 5-HT1A | −0.079 to 0.050 | 0 | 0 | 0 | 0 |

| System | Partial *r* (three constructions) | Moran cells (of 3) | Variogram cells (of 3) | Distribution cells (of 3) | Cells of nine |
|---|---|---|---|---|---|
| 5-HT1B | −0.026 to 0.001 | 0 | 0 | 0 | 0 |
| 5-HT2A | 0.159 to 0.175 | 0 | 0 | 0 | 0 |
| 5-HT4 | 0.056 to 0.137 | 0 | 0 | 0 | 0 |
| 5-HT6 | −0.109 to −0.072 | 0 | 0 | 0 | 0 |
| 5-HTT | −0.352 to −0.176 | 1 | 1 | 1 | 3 |
| CB1 | −0.232 to −0.123 | 1 | 0 | 0 | 1 |
| D1 | −0.266 to −0.006 | 1 | 1 | 1 | 3 |
| D2 | −0.178 to 0.011 | 0 | 0 | 0 | 0 |
| DAT | −0.324 to −0.071 | 1 | 1 | 1 | 3 |
| GABAA | −0.018 to 0.011 | 0 | 0 | 0 | 0 |
| H3 | −0.471 to −0.346 | 3 | 3 | 3 | 9 |
| M1 | −0.065 to −0.017 | 0 | 0 | 0 | 0 |
| mGluR5 | −0.207 to −0.148 | 0 | 0 | 0 | 0 |
| MOR | −0.391 to −0.221 | 1 | 1 | 1 | 3 |
| NET | −0.271 to −0.193 | 0 | 0 | 0 | 0 |
| NMDA | −0.276 to −0.170 | 3 | 2 | 2 | 7 |
| VAChT | −0.509 to −0.319 | 3 | 3 | 3 | 9 |
| α4β2 | −0.224 to −0.212 | 0 | 0 | 0 | 0 |

**Supplementary Table 21. Prespecified tests of the reading-skill map in 169 children (OpenNeuro ds001894).** False discovery control across the three prespecified maps (3) and across the nineteen maps (19).

| Definition | System | *r* | Moran *P* | Moran *q* (3) | Moran *q* (19) | Variogram *P* | Variogram *q* (3) | Distribution *P* | Distribution *q* (3) |
|---|---|---|---|---|---|---|---|---|---|
| Affine plus SyN | H3 | −0.365 | 0.0388 | 0.1164 | 0.2457 | 0.1096 | 0.1644 | 0.1068 | 0.1602 |
| Affine plus SyN | NMDA | −0.145 | 0.0894 | 0.1341 | 0.2665 | 0.0564 | 0.1644 | 0.0584 | 0.1602 |
| Affine plus SyN | VAChT | −0.074 | 0.7550 | 0.7550 | 0.7970 | 0.6519 | 0.6519 | 0.6563 | 0.6563 |
| Nonlinear only | H3 | −0.343 | 0.0234 | 0.0702 | 0.2792 | 0.1090 | 0.3269 | 0.1116 | 0.3347 |
| Nonlinear only | NMDA | −0.046 | 0.5863 | 0.6777 | 0.7574 | 0.5671 | 0.6179 | 0.5589 | 0.6249 |
| Nonlinear only | VAChT | −0.074 | 0.6777 | 0.6777 | 0.7574 | 0.6179 | 0.6179 | 0.6249 | 0.6249 |

**Supplementary Table 22. Sensitivity models for the reading-skill map in 169 children.** Map correlation is the correlation of each alternative map with the primary map of the same definition.

| Definition | Model | *n* | Map correlation with primary | H3 *r* | H3 Moran *P* | NMDA *r* | VAChT *r* |
|---|---|---|---|---|---|---|---|

| Definition | Model | *n* | Map correlation with primary | H3 *r* | H3 Moran *P* | NMDA *r* | VAChT *r* |
|---|---|---|---|---|---|---|---|
| Affine plus SyN | Primary model | 169 | 1.000 | −0.365 | 0.0388 | −0.145 | −0.074 |
| Affine plus SyN | WJ-III Broad Reading in place of TOWRE | 167 | 0.906 | −0.360 | 0.0382 | −0.079 | −0.039 |
| Affine plus SyN | Full-scale IQ added | 169 | 0.596 | −0.148 | 0.0852 | −0.134 | −0.040 |
| Affine plus SyN | MRI quality metrics added | 169 | 0.993 | −0.360 | 0.0264 | −0.137 | −0.046 |
| Affine plus SyN | Global covariate omitted | 169 | 0.971 | −0.353 | 0.0794 | −0.150 | −0.093 |
| Affine plus SyN | Quadratic age added | 169 | 0.999 | −0.358 | 0.0556 | −0.135 | −0.065 |
| Affine plus SyN | Registration and image-quality outliers removed (MAD rule over six quality variables) | 161 | 0.994 | −0.366 | 0.0410 | −0.143 | −0.067 |
| Nonlinear only | Primary model | 169 | 1.000 | −0.343 | 0.0234 | −0.046 | −0.074 |
| Nonlinear only | WJ-III Broad Reading in place of TOWRE | 167 | 0.848 | −0.291 | 0.0634 | 0.047 | −0.017 |
| Nonlinear only | Full-scale IQ added | 169 | 0.596 | −0.103 | 0.3591 | −0.078 | −0.036 |
| Nonlinear only | MRI quality metrics added | 169 | 0.988 | −0.348 | 0.0072 | −0.062 | −0.045 |
| Nonlinear only | Global covariate omitted | 169 | 0.970 | −0.376 | 0.0274 | −0.089 | −0.109 |
| Nonlinear only | Quadratic age added | 169 | 0.997 | −0.333 | 0.0404 | −0.032 | −0.060 |
| Nonlinear only | Registration and image-quality outliers removed (MAD rule over six quality variables) | 161 | 0.991 | −0.357 | 0.0222 | −0.046 | −0.076 |

**Supplementary Table 23. Similarity of the reading-skill map of 169 children to the three adult liability maps.** False discovery control across the three tests.

| Liability map | *r* | Moran *P* | Moran *q* |
|---|---|---|---|
| Unadjusted log-Jacobian | 0.266 | 0.3055 | 0.5497 |
| Raw Jacobian, head size | 0.193 | 0.5497 | 0.5497 |
| Log-Jacobian, head size and education | 0.248 | 0.5115 | 0.5497 |

**Supplementary Table 24. Registration threshold sensitivity in the 169-children analysis.** The primary threshold (0.55) and 0.60 retain the same 169 children because the distribution of warped-template correlation has no scan between 0.525 and 0.633; 0.66 retains 152. Rerun on the analysis cluster with the primary model and seeds.

| Warped-template threshold | *n* | System | *r* | Moran *P* | Variogram *P* | Distribution *P* |
|---|---|---|---|---|---|---|
| 0.55 (primary) | 169 | H3 | −0.365 | 0.0388 | 0.1096 | 0.1068 |
| 0.55 (primary) | 169 | NMDA | −0.145 | 0.0894 | 0.0564 | 0.0584 |
| 0.55 (primary) | 169 | VAChT | −0.074 | 0.7550 | 0.6519 | 0.6563 |
| 0.60 | 169 | H3 | −0.365 | 0.0388 | 0.1096 | 0.1068 |
| 0.60 | 169 | NMDA | −0.145 | 0.0894 | 0.0564 | 0.0584 |
| 0.60 | 169 | VAChT | −0.074 | 0.7550 | 0.6519 | 0.6563 |
| 0.66 | 152 | H3 | −0.316 | 0.1592 | 0.1530 | 0.1520 |
| 0.66 | 152 | NMDA | −0.099 | 0.2559 | 0.2208 | 0.2214 |
| 0.66 | 152 | VAChT | −0.041 | 0.8616 | 0.8020 | 0.8040 |

**Supplementary Table 25. Screen of the published reading-skill maps of 9 013 children against the nineteen maps.** Maps from Carrion-Castillo et al. (2023) over the 74 left-hemisphere Destrieux regions, with the nineteen maps rebuilt at the same regions. False discovery control across the 38 tests.

| Measure | System | *r* | Moran *P* | Moran *q* | Variogram *P* | Variogram *q* | Distribution *P* | Distribution *q* |
|---|---|---|---|---|---|---|---|---|
| Surface area, adjusted | 5-HT1A | 0.224 | 0.7233 | 0.9665 | 0.5539 | 0.9796 | 0.5127 | 0.9754 |
| Surface area, adjusted | 5-HT1B | −0.171 | 0.4127 | 0.9665 | 0.3917 | 0.9796 | 0.3833 | 0.9754 |
| Surface area, adjusted | 5-HT2A | 0.154 | 0.8558 | 0.9665 | 0.5969 | 0.9796 | 0.5807 | 0.9754 |
| Surface area, adjusted | 5-HT4 | 0.228 | 0.8212 | 0.9665 | 0.5045 | 0.9796 | 0.4791 | 0.9754 |
| Surface area, adjusted | 5-HT6 | 0.026 | 0.8728 | 0.9665 | 0.8470 | 0.9796 | 0.8398 | 0.9754 |
| Surface area, adjusted | 5-HTT | 0.110 | 0.8872 | 0.9665 | 0.7389 | 0.9796 | 0.7095 | 0.9754 |
| Surface area, adjusted | CB1 | 0.068 | 0.5655 | 0.9665 | 0.8432 | 0.9796 | 0.8304 | 0.9754 |
| Surface area, adjusted | D1 | −0.153 | 0.1914 | 0.9090 | 0.6289 | 0.9796 | 0.5945 | 0.9754 |
| Surface area, adjusted | D2 | 0.181 | 0.8106 | 0.9665 | 0.6115 | 0.9796 | 0.5783 | 0.9754 |
| Surface area, adjusted | DAT | 0.053 | 0.8080 | 0.9665 | 0.8618 | 0.9796 | 0.8468 | 0.9754 |
| Surface area, adjusted | GABAA | 0.065 | 0.7171 | 0.9665 | 0.8110 | 0.9796 | 0.7976 | 0.9754 |
| Surface area, adjusted | H3 | −0.172 | 0.1546 | 0.9090 | 0.6017 | 0.9796 | 0.5967 | 0.9754 |
| Surface area, adjusted | M1 | −0.202 | 0.0420 | 0.9090 | 0.0980 | 0.9796 | 0.0934 | 0.9754 |
| Surface area, adjusted | mGluR5 | −0.090 | 0.4789 | 0.9665 | 0.6799 | 0.9796 | 0.6575 | 0.9754 |
| Surface area, adjusted | MOR | 0.068 | 0.6129 | 0.9665 | 0.8986 | 0.9796 | 0.8876 | 0.9754 |
| Surface area, adjusted | NET | 0.025 | 0.8336 | 0.9665 | 0.9192 | 0.9796 | 0.9160 | 0.9754 |
| Surface area, adjusted | NMDA | 0.009 | 0.9700 | 0.9796 | 0.9586 | 0.9796 | 0.9652 | 0.9754 |
| Surface area, adjusted | VAChT | 0.183 | 0.3623 | 0.9665 | 0.6137 | 0.9796 | 0.5809 | 0.9754 |
| Surface area, adjusted | α4β2 | 0.062 | 0.6687 | 0.9665 | 0.7712 | 0.9796 | 0.7489 | 0.9754 |
| Thickness, adjusted | 5-HT1A | 0.052 | 0.7950 | 0.9665 | 0.8524 | 0.9796 | 0.8458 | 0.9754 |

| Measure | System | *r* | Moran *P* | Moran *q* | Variogram *P* | Variogram *q* | Distribution *P* | Distribution *q* |
|---|---|---|---|---|---|---|---|---|
| Thickness, adjusted | 5-HT1B | −0.205 | 0.3251 | 0.9665 | 0.2084 | 0.9796 | 0.2092 | 0.9754 |
| Thickness, adjusted | 5-HT2A | 0.036 | 0.8642 | 0.9665 | 0.8666 | 0.9796 | 0.8684 | 0.9754 |
| Thickness, adjusted | 5-HT4 | 0.107 | 0.6739 | 0.9665 | 0.6777 | 0.9796 | 0.6761 | 0.9754 |
| Thickness, adjusted | 5-HT6 | −0.041 | 0.8902 | 0.9665 | 0.7213 | 0.9796 | 0.7357 | 0.9754 |
| Thickness, adjusted | 5-HTT | 0.300 | 0.1894 | 0.9090 | 0.1448 | 0.9796 | 0.1372 | 0.9754 |
| Thickness, adjusted | CB1 | −0.066 | 0.6751 | 0.9665 | 0.7930 | 0.9796 | 0.7854 | 0.9754 |
| Thickness, adjusted | D1 | −0.015 | 0.9316 | 0.9796 | 0.9490 | 0.9796 | 0.9520 | 0.9754 |
| Thickness, adjusted | D2 | 0.193 | 0.3511 | 0.9665 | 0.4425 | 0.9796 | 0.4325 | 0.9754 |
| Thickness, adjusted | DAT | 0.164 | 0.3659 | 0.9665 | 0.4371 | 0.9796 | 0.4173 | 0.9754 |
| Thickness, adjusted | GABAA | 0.220 | 0.1258 | 0.9090 | 0.2579 | 0.9796 | 0.2569 | 0.9754 |
| Thickness, adjusted | H3 | −0.179 | 0.1756 | 0.9090 | 0.4761 | 0.9796 | 0.4543 | 0.9754 |
| Thickness, adjusted | M1 | 0.060 | 0.6409 | 0.9665 | 0.6295 | 0.9796 | 0.6263 | 0.9754 |
| Thickness, adjusted | mGluR5 | −0.185 | 0.5383 | 0.9665 | 0.2484 | 0.9796 | 0.2474 | 0.9754 |
| Thickness, adjusted | MOR | −0.324 | 0.1428 | 0.9090 | 0.3775 | 0.9796 | 0.3627 | 0.9754 |
| Thickness, adjusted | NET | 0.005 | 0.9796 | 0.9796 | 0.9796 | 0.9796 | 0.9754 | 0.9754 |
| Thickness, adjusted | NMDA | −0.023 | 0.8664 | 0.9665 | 0.8746 | 0.9796 | 0.8844 | 0.9754 |
| Thickness, adjusted | VAChT | −0.073 | 0.7155 | 0.9665 | 0.7856 | 0.9796 | 0.7842 | 0.9754 |
| Thickness, adjusted | α4β2 | −0.258 | 0.1506 | 0.9090 | 0.1024 | 0.9796 | 0.0992 | 0.9754 |

**Supplementary Table 26. Similarity of the published reading-skill maps of 9 013 children to the three adult liability maps.** False discovery control across the six tests.

| Skill map | Liability map | *r* | Moran *P* | Moran *q* | Variogram *q* | Distribution *q* |
|---|---|---|---|---|---|---|
| Surface area, adjusted | Unadjusted log-Jacobian | −0.057 | 0.6241 | 0.8442 | 0.9378 | 0.9420 |
| Surface area, adjusted | Raw Jacobian, head size | −0.044 | 0.7035 | 0.8442 | 0.9378 | 0.9420 |
| Surface area, adjusted | Log-Jacobian, head size and education | −0.018 | 0.8780 | 0.8780 | 0.9378 | 0.9420 |
| Thickness, adjusted | Unadjusted log-Jacobian | 0.209 | 0.2362 | 0.8442 | 0.9274 | 0.9046 |
| Thickness, adjusted | Raw Jacobian, head size | 0.136 | 0.4233 | 0.8442 | 0.9274 | 0.9046 |
| Thickness, adjusted | Log-Jacobian, head size and education | 0.183 | 0.3117 | 0.8442 | 0.9274 | 0.9046 |

**Supplementary Table 27. Formal comparison of the adult liability and child skill correlations with H3, reliability and power.** Williams tests treat the adult and child correlations with H3 as dependent through the correlation between the two maps. Reliability is one minus the mean squared regional standard error over the variance of the regional estimates; disattenuation divides the child correlation by the square root of reliability. Power is computed at the corrected two-sided threshold of the prespecified family from the Moran null distribution of the child map, for the weakest adult H3 correlation (−0.475) and for the child map's own correlation.

| Sample | Adult construction | *r* adult, H3 | *r* child, H3 | *r* adult, child | Williams *t* (218) | *P* | Williams *t* (39) | *P* | Child map reliability | Child *r* disattenuated | Power, weakest adult *r* | Power, own *r* |
|---|---|---|---|---|---|---|---|---|---|---|---|---|
| 169 children (ds001894) | Unadjusted log-Jacobian | −0.618 | −0.365 | 0.266 | -3.86 | 0.0001 | -1.58 | 0.1222 | 0.48 | −0.527 | 0.54 | 0.18 |
| 169 children (ds001894) | Raw Jacobian, head size | −0.475 | −0.365 | 0.193 | -1.47 | 0.1430 | -0.60 | 0.5507 | 0.48 | −0.527 | 0.32 | 0.18 |
| 169 children (ds001894) | Log-Jacobian, head size and education | −0.481 | −0.365 | 0.248 | -1.60 | 0.1103 | -0.66 | 0.5155 | 0.48 | −0.527 | 0.33 | 0.18 |

**Supplementary Table 28. Per-person neuromodulator alignment and reading skill in 169 children (prespecified 6 September 2026).** Each child's 218-region deviation vector (regional log-Jacobian residualised across children on age, sex and mean brain log-Jacobian, then standardised) was correlated with the H3, VAChT and NMDA maps and with the shared axis. Reading score was regressed on each score with the same covariates (HC3 standard errors; 5 000 subject permutations); false discovery control across the four scores. The prespecified direction was a positive coefficient; the observed coefficients are negative.

| Specification | *n* | Map | β (TOWRE points per SD of score) | 95% interval | HC3 *P* | Permutation *P* | *q* (4) | Score *r* with global log-Jacobian |
|---|---|---|---|---|---|---|---|---|
| primary | 169 | H3 | −3.41 | −5.90 to −0.93 | 0.0078 | 0.0098 | 0.0311 | −0.01 |
| primary | 169 | VAChT | −0.80 | −3.59 to 1.99 | 0.5733 | 0.5725 | 0.5733 | 0.00 |
| primary | 169 | NMDA | −1.93 | −4.47 to 0.61 | 0.1374 | 0.1372 | 0.1833 | 0.01 |
| primary | 169 | shared axis | −3.10 | −5.83 to −0.38 | 0.0270 | 0.0316 | 0.0540 | 0.00 |
| nonlinear-only definition | 169 | H3 | −3.44 | −5.92 to −0.96 | 0.0073 | 0.0060 | 0.0292 | −0.01 |
| nonlinear-only definition | 169 | VAChT | −0.83 | −3.63 to 1.96 | 0.5601 | 0.5511 | 0.5601 | 0.00 |
| nonlinear-only definition | 169 | NMDA | −1.88 | −4.42 to 0.66 | 0.1480 | 0.1504 | 0.1973 | 0.01 |
| nonlinear-only definition | 169 | shared axis | −3.10 | −5.82 to −0.37 | 0.0273 | 0.0236 | 0.0545 | 0.00 |
| WJ-III Broad Reading | 167 | H3 | −2.69 | −4.76 to −0.61 | 0.0120 | 0.0104 | 0.0480 | −0.01 |
| WJ-III Broad Reading | 167 | VAChT | −0.12 | −2.28 to 2.04 | 0.9140 | 0.9066 | 0.9140 | 0.00 |
| WJ-III Broad Reading | 167 | NMDA | −0.83 | −2.99 to 1.33 | 0.4538 | 0.4499 | 0.6051 | 0.01 |
| WJ-III Broad Reading | 167 | shared axis | −1.74 | −4.06 to 0.57 | 0.1418 | 0.1418 | 0.2836 | 0.00 |
| quadratic age | 169 | H3 | −3.40 | −5.87 to −0.93 | 0.0076 | 0.0074 | 0.0306 | −0.01 |
| quadratic age | 169 | VAChT | −0.68 | −3.51 to 2.15 | 0.6370 | 0.6273 | 0.6370 | 0.00 |
| quadratic age | 169 | NMDA | −1.79 | −4.28 to 0.70 | 0.1606 | 0.1524 | 0.2142 | 0.01 |
| quadratic age | 169 | shared axis | −3.09 | −5.79 to −0.39 | 0.0264 | 0.0284 | 0.0528 | 0.00 |
| registration threshold 0.66 | 152 | H3 | −2.85 | −5.40 to −0.30 | 0.0298 | 0.0298 | 0.0878 | −0.01 |
| registration threshold 0.66 | 152 | VAChT | −0.35 | −3.27 to 2.58 | 0.8163 | 0.7970 | 0.8163 | 0.00 |
| registration threshold 0.66 | 152 | NMDA | −1.23 | −3.87 to 1.41 | 0.3626 | 0.3597 | 0.4834 | 0.02 |
| registration threshold 0.66 | 152 | shared axis | −2.93 | −5.75 to −0.10 | 0.0439 | 0.0412 | 0.0878 | 0.00 |

**Supplementary Table 29. Liability constructions against three maps of dyslexia itself.** Case-control functional and structural syntheses and the task-based fMRI group map, with false discovery control across the nine tests.

| Dyslexia map | Construction | *r* | Moran *q* | Variogram *q* | Distribution *q* |
|---|---|---|---|---|---|
| Case-control functional synthesis | Unadjusted log-Jacobian | 0.139 | 0.6337 | 0.7178 | 0.6998 |
| Case-control functional synthesis | Raw Jacobian, head size | 0.052 | 0.6615 | 0.7738 | 0.7822 |
| Case-control functional synthesis | Log-Jacobian, head size and education | 0.086 | 0.6337 | 0.7302 | 0.7147 |
| Case-control structural synthesis | Unadjusted log-Jacobian | 0.091 | 0.6337 | 0.7178 | 0.6998 |
| Case-control structural synthesis | Raw Jacobian, head size | 0.065 | 0.6436 | 0.7302 | 0.7147 |
| Case-control structural synthesis | Log-Jacobian, head size and education | 0.102 | 0.6337 | 0.7178 | 0.6998 |
| Task-fMRI group map | Unadjusted log-Jacobian | 0.149 | 0.6337 | 0.7178 | 0.6998 |
| Task-fMRI group map | Raw Jacobian, head size | 0.145 | 0.6337 | 0.7178 | 0.6998 |
| Task-fMRI group map | Log-Jacobian, head size and education | 0.130 | 0.6337 | 0.7178 | 0.6998 |

**Supplementary Table 30. Source-map uncertainty for the six systems with matched source images.** Source sample is the number of participants behind the source image. Agreement is the correlation between the source reconstruction and its atlas counterpart. Constructions reaching $q < 0.05$ are counted of three under each procedure. Sign stability is the share of 5 000 perturbations in which the correlation kept its sign.

| System | Source sample | Agreement with atlas, *r* | Source-mean *r* | Moran | Variogram | Distribution | Sign stability |
|---|---|---|---|---|---|---|---|
| 5-HT2A | 29 | 0.728 | 0.169 to 0.233 | 0 of 3 | 0 of 3 | 0 of 3 | 100.0% |
| 5-HT4 | 59 | 0.607 | 0.045 to 0.199 | 0 of 3 | 0 of 3 | 0 of 3 | 100.0% |
| 5-HTT | 100 | 0.868 | −0.172 to 0.013 | 0 of 3 | 0 of 3 | 0 of 3 | 100.0% |
| GABAA | 16 | 0.594 | 0.155 to 0.225 | 0 of 3 | 0 of 3 | 0 of 3 | 100.0% |
| NMDA | 29 | 0.761 | −0.310 to −0.135 | 0 of 3 | 1 of 3 | 1 of 3 | 99.5% |
| mGluR5 | 100 | 0.989 | −0.375 to −0.309 | 0 of 3 | 1 of 3 | 3 of 3 | 100.0% |

**Supplementary Table 31. Prespecified tests of the external genetic comparison (GenLang x ENIGMA3-only).** Regional genetic correlations over 34 bilateral Desikan-Killiany regions, with false discovery control across the six tests. Correlations are oriented to dyslexia liability, the reading-oriented genetic correlations having been multiplied by −1 before testing, so negative values have the same sense as the main analysis.

| Measure | System | *r* | Moran *P* | Moran *q* | Variogram *P* | Variogram *q* | Distribution *P* | Distribution *q* |
|---|---|---|---|---|---|---|---|---|
| Surface area | H3 | −0.216 | 0.3591 | 0.7183 | 0.4909 | 0.7379 | 0.4833 | 0.7460 |
| Surface area | NMDA | −0.231 | 0.2028 | 0.6083 | 0.1372 | 0.6353 | 0.1442 | 0.5987 |

| Measure | System | *r* | Moran *P* | Moran *q* | Variogram *P* | Variogram *q* | Distribution *P* | Distribution *q* |
|---|---|---|---|---|---|---|---|---|
| Surface area | VAChT | −0.288 | 0.1292 | 0.6083 | 0.2118 | 0.6353 | 0.1996 | 0.5987 |
| Thickness | H3 | 0.143 | 0.6261 | 0.7513 | 0.7041 | 0.7379 | 0.6959 | 0.7568 |
| Thickness | NMDA | 0.049 | 0.8882 | 0.8882 | 0.7379 | 0.7379 | 0.7568 | 0.7568 |
| Thickness | VAChT | −0.186 | 0.5223 | 0.7513 | 0.5089 | 0.7379 | 0.4973 | 0.7460 |

**Supplementary Table 32. Full 38-test external genetic screen (with-global models).** Same orientation as the previous table; false discovery control across the 38 tests within each procedure.

| Measure | System | *r* | Moran *P* | Moran *q* | Variogram *P* | Variogram *q* | Distribution *P* | Distribution *q* |
|---|---|---|---|---|---|---|---|---|
| Surface area | 5-HT1A | −0.461 | 0.1642 | 0.7798 | 0.0994 | 0.8553 | 0.0894 | 0.8461 |
| Surface area | 5-HT1B | 0.264 | 0.1608 | 0.7798 | 0.1374 | 0.8553 | 0.1442 | 0.8461 |
| Surface area | 5-HT2A | 0.174 | 0.5219 | 0.9462 | 0.4153 | 0.8553 | 0.4061 | 0.8461 |
| Surface area | 5-HT4 | −0.123 | 0.7996 | 0.9640 | 0.6575 | 0.9609 | 0.6363 | 0.9299 |
| Surface area | 5-HT6 | 0.003 | 0.9834 | 0.9834 | 0.9862 | 0.9862 | 0.9842 | 0.9842 |
| Surface area | 5-HTT | −0.246 | 0.4191 | 0.9368 | 0.2561 | 0.8553 | 0.2523 | 0.8461 |
| Surface area | CB1 | −0.314 | 0.1628 | 0.7798 | 0.3541 | 0.8553 | 0.3371 | 0.8461 |
| Surface area | D1 | −0.226 | 0.2398 | 0.8282 | 0.2671 | 0.8553 | 0.2535 | 0.8461 |
| Surface area | D2 | −0.560 | 0.0108 | 0.2052 | 0.0264 | 0.5015 | 0.0188 | 0.3571 |
| Surface area | DAT | −0.596 | 0.0284 | 0.3597 | 0.0008 | 0.0304 | 0.0006 | 0.0228 |
| Surface area | GABAA | 0.415 | 0.0062 | 0.2052 | 0.0420 | 0.5319 | 0.0402 | 0.5091 |
| Surface area | H3 | −0.216 | 0.3591 | 0.9368 | 0.4909 | 0.8553 | 0.4833 | 0.8461 |
| Surface area | M1 | 0.134 | 0.3099 | 0.9368 | 0.3841 | 0.8553 | 0.3921 | 0.8461 |
| Surface area | mGluR5 | −0.106 | 0.4041 | 0.9368 | 0.5913 | 0.9070 | 0.5705 | 0.8790 |
| Surface area | MOR | −0.372 | 0.0434 | 0.4122 | 0.3081 | 0.8553 | 0.2971 | 0.8461 |
| Surface area | NET | 0.125 | 0.5229 | 0.9462 | 0.5967 | 0.9070 | 0.5783 | 0.8790 |
| Surface area | NMDA | −0.231 | 0.2028 | 0.8282 | 0.1372 | 0.8553 | 0.1442 | 0.8461 |
| Surface area | VAChT | −0.288 | 0.1292 | 0.7798 | 0.2118 | 0.8553 | 0.1996 | 0.8461 |
| Surface area | α4β2 | 0.204 | 0.3287 | 0.9368 | 0.3243 | 0.8553 | 0.3175 | 0.8461 |
| Thickness | 5-HT1A | −0.084 | 0.6911 | 0.9640 | 0.8208 | 0.9652 | 0.8152 | 0.9662 |
| Thickness | 5-HT1B | 0.026 | 0.9104 | 0.9640 | 0.9020 | 0.9793 | 0.8936 | 0.9702 |
| Thickness | 5-HT2A | −0.159 | 0.4589 | 0.9462 | 0.5177 | 0.8553 | 0.5085 | 0.8461 |
| Thickness | 5-HT4 | −0.214 | 0.2232 | 0.8282 | 0.5137 | 0.8553 | 0.5121 | 0.8461 |
| Thickness | 5-HT6 | 0.097 | 0.7984 | 0.9640 | 0.4849 | 0.8553 | 0.4947 | 0.8461 |
| Thickness | 5-HTT | −0.261 | 0.4165 | 0.9368 | 0.3233 | 0.8553 | 0.3157 | 0.8461 |
| Thickness | CB1 | 0.023 | 0.9132 | 0.9640 | 0.9610 | 0.9862 | 0.9530 | 0.9842 |
| Thickness | D1 | 0.275 | 0.5537 | 0.9564 | 0.2124 | 0.8553 | 0.2028 | 0.8461 |
| Thickness | D2 | −0.100 | 0.6441 | 0.9640 | 0.7762 | 0.9652 | 0.7826 | 0.9662 |

| **Measure** | **System** | ***r*** | **Moran *P*** | **Moran *q*** | **Variogram *P*** | **Variogram *q*** | **Distribution *P*** | **Distribution *q*** |
|---|---|---|---|---|---|---|---|---|
| Thickness | DAT | −0.052 | 0.7894 | 0.9640 | 0.8362 | 0.9652 | 0.8390 | 0.9662 |
| Thickness | GABAA | −0.056 | 0.8406 | 0.9640 | 0.8382 | 0.9652 | 0.8342 | 0.9662 |
| Thickness | H3 | 0.143 | 0.6261 | 0.9640 | 0.7041 | 0.9652 | 0.6959 | 0.9662 |
| Thickness | M1 | 0.171 | 0.7287 | 0.9640 | 0.2396 | 0.8553 | 0.2320 | 0.8461 |
| Thickness | mGluR5 | 0.149 | 0.9102 | 0.9640 | 0.4877 | 0.8553 | 0.4819 | 0.8461 |
| Thickness | MOR | 0.015 | 0.9446 | 0.9701 | 0.9720 | 0.9862 | 0.9734 | 0.9842 |
| Thickness | NET | −0.081 | 0.8080 | 0.9640 | 0.7632 | 0.9652 | 0.7584 | 0.9662 |
| Thickness | NMDA | 0.049 | 0.8882 | 0.9640 | 0.7379 | 0.9652 | 0.7568 | 0.9662 |
| Thickness | VAChT | −0.186 | 0.5223 | 0.9462 | 0.5089 | 0.8553 | 0.4973 | 0.8461 |
| Thickness | α4β2 | −0.046 | 0.8768 | 0.9640 | 0.8708 | 0.9733 | 0.8648 | 0.9666 |

**Supplementary Table 33. Exploratory triangulation with 22 preselected UK Biobank language regions.** Multivariate GenLang genetic correlations with 22 preselected imaging traits, in the reading orientation. The imaging side is the parent UK Biobank cohort and the mask is preselected, so this family is exploratory. False discovery control across the six tests.

| **Measure** | **System** | ***r*** | **Moran *P*** | **Moran *q* (6)** | **Spearman *r*** | **Moran *P*** | **Inverse-variance *r*** | **Moran *P*** | **Leave-one-pair *r*** |
|---|---|---|---|---|---|---|---|---|---|
| Surface area | H3 | −0.282 | 0.2144 | 0.4287 | −0.151 | 0.4517 | −0.245 | 0.2933 | −0.396 to −0.082 |
| Surface area | NMDA | −0.253 | 0.3695 | 0.5478 | −0.238 | 0.4365 | −0.294 | 0.3165 | −0.383 to 0.017 |
| Surface area | VAChT | −0.246 | 0.4565 | 0.5478 | −0.240 | 0.5441 | −0.265 | 0.4317 | −0.301 to −0.112 |
| Thickness | H3 | −0.017 | 0.9548 | 0.9548 | 0.029 | 0.9280 | 0.004 | 0.9906 | −0.287 to 0.163 |
| Thickness | NMDA | 0.487 | 0.0818 | 0.2454 | 0.455 | 0.1466 | 0.473 | 0.0712 | 0.286 to 0.602 |
| Thickness | VAChT | 0.622 | 0.0058 | 0.0348 | 0.470 | 0.0754 | 0.631 | 0.0034 | 0.427 to 0.674 |

**Supplementary Table 34. Stratified phenotype family (Family C).** Liability constructions against case-control synthesis strata, with false discovery control across the 18 tests within each procedure; prespecified in an addendum that recorded the override of the parent document.

| **Construction** | **Phenotype stratum** | ***r*** | **Moran *q*** | **Variogram *q*** | **Distribution *q*** |
|---|---|---|---|---|---|
| Unadjusted log-Jacobian | structural reduction only | 0.084 | 0.9414 | 0.9572 | 0.9526 |
| Unadjusted log-Jacobian | structural increase only | −0.022 | 0.9414 | 0.9572 | 0.9526 |
| Unadjusted log-Jacobian | structural adult ge216m | 0.081 | 0.9414 | 0.9572 | 0.9526 |
| Unadjusted log-Jacobian | structural pediatric lt216m | 0.057 | 0.9414 | 0.9572 | 0.9526 |
| Unadjusted log-Jacobian | functional adults | 0.103 | 0.9414 | 0.9572 | 0.9526 |
| Unadjusted log-Jacobian | functional children | 0.090 | 0.9414 | 0.9572 | 0.9526 |
| Raw Jacobian, head size | structural reduction only | 0.041 | 0.9414 | 0.9572 | 0.9526 |
| Raw Jacobian, head size | structural increase only | −0.047 | 0.9414 | 0.9572 | 0.9526 |

| Construction | Phenotype stratum | *r* | Moran *q* | Variogram *q* | Distribution *q* |
|---|---|---|---|---|---|
| Raw Jacobian, head size | structural adult ge216m | 0.064 | 0.9414 | 0.9572 | 0.9526 |
| Raw Jacobian, head size | structural pediatric lt216m | 0.038 | 0.9414 | 0.9572 | 0.9526 |
| Raw Jacobian, head size | functional adults | 0.029 | 0.9414 | 0.9646 | 0.9621 |
| Raw Jacobian, head size | functional children | 0.005 | 0.9414 | 0.9720 | 0.9800 |
| Log-Jacobian, head size and education | structural reduction only | 0.085 | 0.9414 | 0.9572 | 0.9526 |
| Log-Jacobian, head size and education | structural increase only | −0.044 | 0.9414 | 0.9572 | 0.9526 |
| Log-Jacobian, head size and education | structural adult ge216m | 0.092 | 0.9414 | 0.9572 | 0.9526 |
| Log-Jacobian, head size and education | structural pediatric lt216m | 0.064 | 0.9414 | 0.9572 | 0.9526 |
| Log-Jacobian, head size and education | functional adults | 0.087 | 0.9414 | 0.9572 | 0.9526 |
| Log-Jacobian, head size and education | functional children | 0.010 | 0.9414 | 0.9720 | 0.9800 |
